# Characterization of a pancreatic cancer GWAS signal suggests PDX1 buffers stress in the exocrine pancreas

**DOI:** 10.64898/2026.04.13.26350790

**Authors:** Jason W. Hoskins, Trevor A. Christensen, Daina Eiser, Erin Char, Michael Mobaraki, Aidan O’Brien, Irene Collins, Jun Zhong, Minal B. Patel, Gauri Prasad, Pancreatic Cancer Cohort Consortium and Pancreatic Cancer Case-Control Consortium (PanScan/PanC4), H. Efsun Arda, Katelyn E. Connelly, Laufey T. Amundadottir

## Abstract

Pancreatic ductal adenocarcinoma (PDAC) remains one of the deadliest human cancers. The current largest published PDAC Genome-Wide Association Study (GWAS) identified 23 genetic risk signals, but most lack sufficient characterization. This study aimed to functionally characterize the chr13q12.2 (*PLUT*/*PDX1*) PDAC GWAS risk locus. Fine-mapping, luciferase reporter assays, and electrophoretic mobility shift assays implicated rs9581943, a *PDX1* promoter SNP, as a functional variant underlying this GWAS signal. GTEx expression QTL analyses identified rs9581943 as a significant *PDX1* eQTL in pancreas, and CRISPR/Cas9 editing in PDAC-derived cell lines confirmed a functional relationship. PDX1 is a transcription factor involved in early pancreas development and β-cell homeostasis, but its role in exocrine pancreatic cells is unclear. Single-nucleus RNA-seq analyses of pancreatic acinar and ductal cells from neonatal, adult, and chronic pancreatitis donors suggested PDX1 activity alleviates high secretory load and ER-stress in acinar and biases ducts toward homeostatic phenotypes. Similarly, scRNA-seq analyses of pancreatic tumors suggested PDX1 activity reduces biosynthetic and inflammatory stress and promotes epithelial differentiation. Our study therefore implicates rs9581943 as a causal variant for the chr13q12.2 PDAC GWAS signal wherein the risk allele reduces *PDX1* expression, eroding PDX1’s capacity to buffer stress and stabilize epithelial cell fate in the exocrine compartment.

## Introduction

The biomedical community has made promising strides in the fight against cancer. However, progress in addressing pancreatic cancer has been more modest as demonstrated by a slight increase in its mortality rate in the United States over the past two decades.^1^ Further research into its etiology is critical to reverse this trend. Genome-Wide Association Studies (GWAS) have identified common genetic signals scattered across the genome that are associated with risk of developing pancreatic ductal adenocarcinoma (PDAC), the most common form of pancreatic cancer.^2–7^ Identifying GWAS signals can directly aid in PDAC risk stratification, and investigating the biology underlying these genetic associations can nominate previously unrecognized genes and pathways influencing tumorigenesis that may be susceptible to therapeutic interventions.^8–10^

Investigating germline genetic influences on complex traits, like pancreatic cancer risk, remains an active and promising frontier, but efforts to model the functional consequences of GWAS signals face several challenges.^9,10^ First, germline variants are typically correlated to varying degrees with many others due to linkage disequilibrium (LD), requiring fine-mapping and functional analyses to identify which variant(s) are responsible for a given GWAS signal. Second, the vast majority of significant GWAS variants lie in non-coding genomic regions and do not always affect their nearest genes, and so experiments (e.g., expression QTLs, chromatin looping, or CRISPR-based interventions) are needed to nominate likely effector genes. Third, depending on the GWAS trait, it is often difficult to know *a priori* which cell types/states within a relevant tissue may be mediating the phenotypic effects of a genetic signal, which may require performing the above-mentioned experiments in multiple cell types and/or contexts, perhaps with support from cell type-specific epigenetic regulatory marks. Fourth, even once likely causal variants, effector genes, and cell states are identified, it is often desirable to understand the allele-specific regulatory mechanisms to empower prediction and intervention, which may require a wide range of experiments to elucidate (e.g., electrophoretic mobility shift assays, *in silico* TF-binding predictions, allele-resolved proteomic or epigenomic screens, etc.). Finally, unless an effector gene and its downstream consequences are already well established in the relevant cell state, these must also be modeled experimentally to truly understand the genotype-phenotype relationships of a GWAS signal.

There is still much to learn from the genetic influences on pancreatic cancer risk, but some suggestive patterns have already begun to emerge from published studies.^11–17^ Multiple PDAC GWAS signals have been shown to influence endoplasmic reticulum (ER) stress in acinar cells of the exocrine pancreatic compartment that are responsible for high-throughput production of digestive enzymes.^14,16,17^ Such ER stress can result in chronic inflammation that drives tumorigenesis.^18^ Another suggestive pattern is the presence of several GWAS signals lying in or near genes encoding transcription factors (TFs) with established roles in pancreatic development, regeneration, and homeostasis (e.g., *NR5A2*, *PDX1*, *HNF1A*, *HNF1B*, and *HNF4G*).^7,12,19–24^ Notably, transcriptomic analyses have revealed differential expression and/or activity of some of these TFs in pancreatic tumors compared to normal bulk pancreas^12,25^ or in tumor cell subtypes.^26,27^ This study aimed to functionally model the effects of one such PDAC risk GWAS signal at chr13q12.2 near the gene encoding the master pancreatic TF, *PDX1*. Through fine-mapping, luciferase reporter assays, electrophoretic mobility shift assays, single-base CRISPR/Cas9 editing, and eQTL results, we have identified the *PDX1* promoter SNP rs9581943 as a functional variant underlying this genetic signal that allele-specifically influences the expression of *PDX1*. Overexpressing *PDX1* in pancreatic tumor-derived cell lines resulted in decreased proliferation, likely due to increased apoptosis rates. Consistent with its established role in pancreas development, PDX1 activity was associated with coordinated regulation of exocrine cell-state programs in both normal and malignant tissue. Single-cell analyses of healthy, inflamed, and tumor pancreas suggest that PDX1 acts as a context-dependent buffering factor that suppresses secretory and inflammatory stress responses while maintaining more epithelial and homeostatic cellular phenotypes, suggesting that reduced PDX1 activity may permit the emergence of metaplastic and more aggressive tumor cell states.

## Results

### Fine-mapping the PDAC GWAS signal at the chr13q12.2 *PDX1/PLUT* locus

A meta-analysis of the Pancreatic Cancer Cohort Consortium and the Pancreatic Cancer Case-Control Consortium (PanScan phases I-III and PanC4) GWAS identified a risk association signal at chromosome 13q12.2 tagged by the lead SNP rs2297316 (OR = 1.17 and *P* = 4.43 x 10^-13^; **Figure 1A** and **Table S1**).^7^ Statistical fine-mapping by SuSiE^28,29^ identified a single credible set composed of 21 variants in very high linkage disequilibrium (r^2^ > 0.95; **Figure 1B** and **Table S2**). Overlaying this credible set with previously published epigenomic annotations (ATAC-seq, histone mark ChIP-seq, and ChromHMM chromatin states) from human pancreas-derived cell lines^30^ and purified human acinar, duct, α, and β cells^31^ indicated only rs2297316 and rs9581943 lay in regions with suggestive regulatory potential in pancreatic contexts (**Figure 1C**). Of those two, only rs9581943 lies within an open chromatin region (i.e., an ATAC-seq peak) observed consistently across pancreatic cell contexts at the gene promoter for the pancreatic lineage regulator PDX1 (**Figure 1C**).

**Figure 1:**
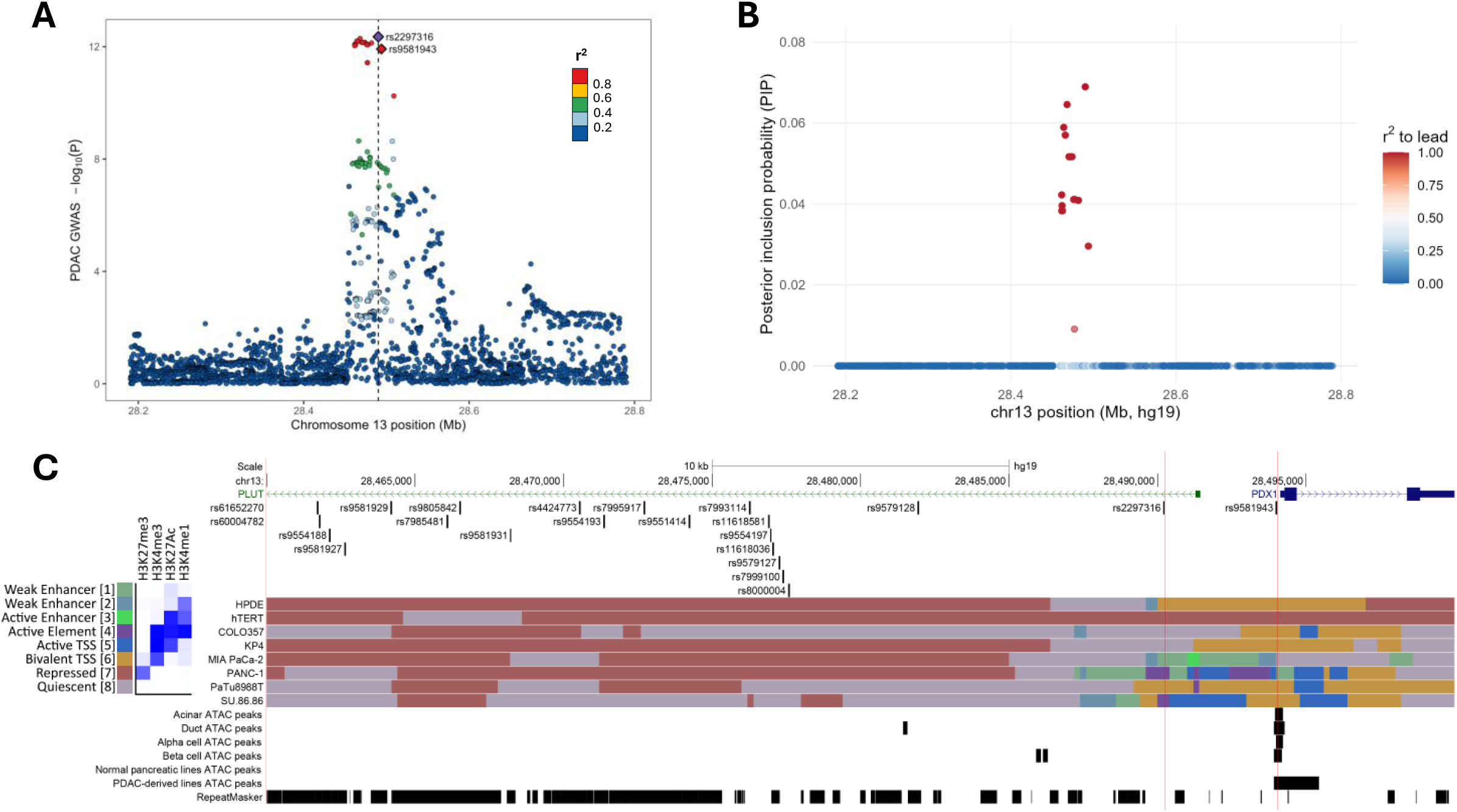
A PDAC GWAS signal at the chr13q12.2 *PDX1*/*PLUT* locus. **(A)** A Manhattan plot of the PanScan (phases I, II, III) and PanC4 meta-analyzed PDAC GWAS results at the chr13q12.2 *PDX1*/*PLUT* locus. SNPs are colored according to their r^2^ with the lead variant, rs2297316. **(B)** Plot of posterior probabilities for SNP inclusion in the 95% credible set of causal variants for the GWAS signal. SNPs are colored according to their r^2^ with the lead variant, rs2297316. **(C)** Genome browser view of the *PDX1*/*PLUT* region overlaying credible set SNPs with pancreatic epigenomic annotations, indicating SNPs with potential regulatory potential. Vertical red lines demonstrate overlap of rs2297316 and rs9581943 with putatively active regulatory regions.

### rs9581943 genotype influences expression regulatory activity

Based on the above statistical fine-mapping and epigenomic annotation overlaps, we prioritized three individual SNPs (rs2297316, rs9581943, rs9579128) and a cluster of seven closely-spaced SNPs (rs7993114, rs11618581, rs9554197, rs11618036, rs9579127, rs7999100, rs8000004) for testing allele-specific regulatory potential by luciferase reporter assays. Constructs were cloned into a reporter vector in both forward and reverse orientations for each allele of each SNP (for the seven SNPs in the cluster, the reference or alternate allele status matched across all seven SNPs), and luciferase reporter assays were performed in both PANC-1 and MIA PaCa-2 pancreatic cell lines (**Figure 2**). Among the rs2297316, rs9579128, and SNP cluster constructs, each had suggestive allele-specific regulatory effects in one of the two orientations in at least one cell line (**Figures 2B-D** and **Table S3**). However, constructs containing rs9581943 demonstrated more significant allele-specific regulatory differences in both orientations and both cell lines (**Figure 2A** and **Table S3**).

**Figure 2:**
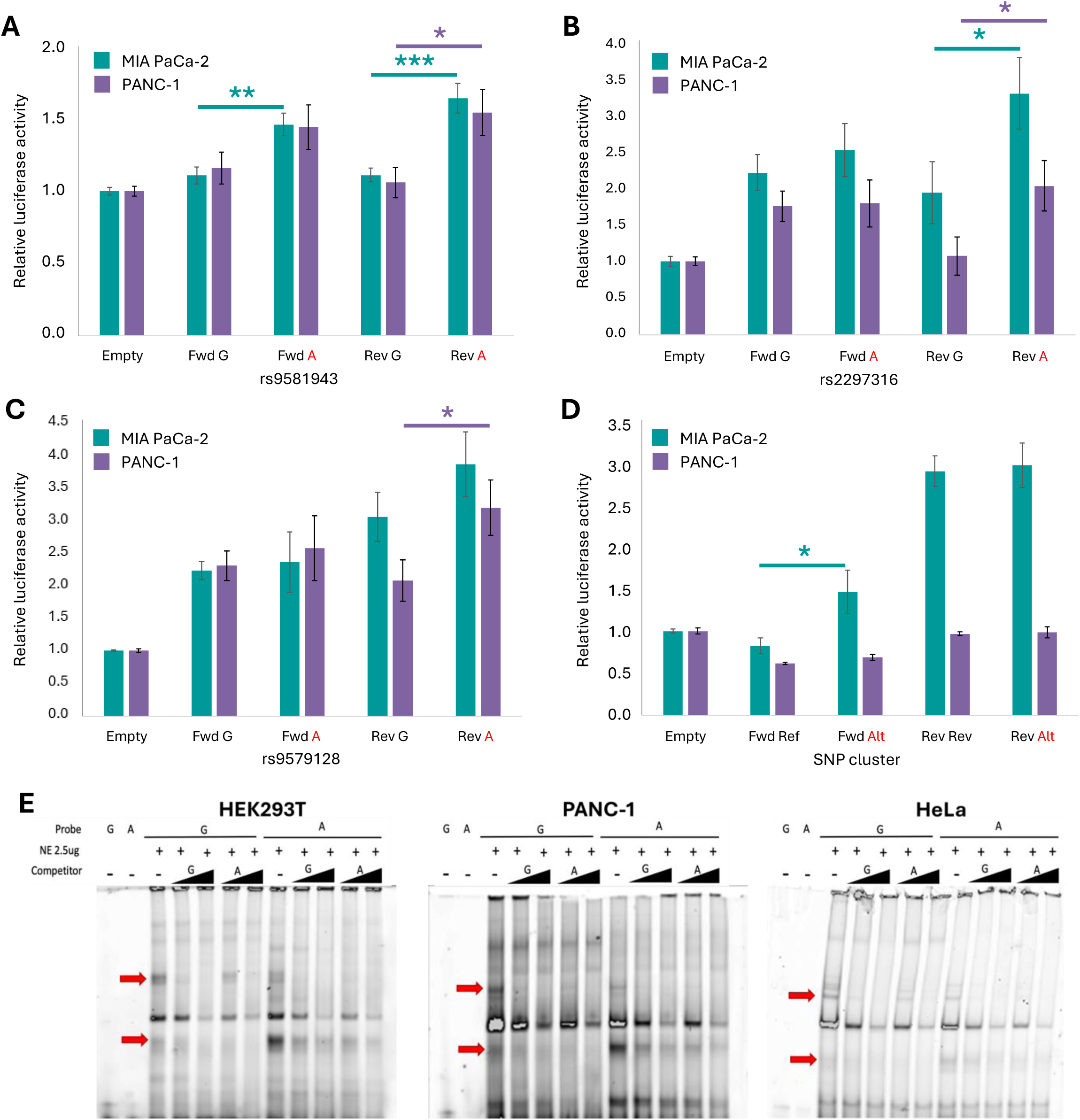
Evidence for rs9581943 allele-specific regulatory activity. **(A-D)** Luciferase reporter assays of rs9581943, rs2297316, rs9579128, and a cluster of seven SNPs (rs7993114, rs11618581, rs9554197, rs11618036, rs9579127, rs7999100, rs8000004), respectively, with their surrounding sequences tested as enhancers upstream of a minimal promoter. Each allele for each SNP (or high-LD SNP cluster) fragment was tested in both forward and reverse orientations in PANC-1 and MIA PaCa-2 cell lines in at least three independent experiments. Error bars represent the standard error of the mean, and *, **, and *** indicate *P* < 0.05, 0.01, and 0.001 by unpaired, two-tailed t-test, respectively. **(E)** Electrophoretic mobility shift assays (EMSA) of rs9581943 major (G) or minor (A) allele probes incubated with 0, 10X, or 50X unlabeled competitor DNA and nuclear extract from HEK293T, PANC-1, or HeLa cell lines. Red arrows indicate G (upper) or A (lower) preferential protein-binding bands.

These results, along with the genomic and epigenomic context at this locus, nominated rs9581943 as the most likely functional variant. Next, we investigated allele-preferential protein binding to dsDNA probes centered on rs9581943 using electrophoretic mobility shift assay (EMSA) with nuclear extracts from three different cell lines (HEK293T, PANC-1, and HeLa). Across all three cell lines, EMSAs revealed both G and A allele-preferential bands, and allelic preference was further demonstrated by competition with each allele’s unlabeled competitor DNA (**Figure 2E**). This suggests allele-preferential protein-binding affinity at this promoter SNP, as expected for a functional regulatory variant.

### rs9581943 underlies a pancreatic *PDX1* eQTL signal colocalizing with the GWAS signal

Given its position merely 140bp upstream of the *PDX1* transcription start site (TSS) and the above evidence of allele-preferential regulatory activity, the observation that rs9581943 (along with highly correlated variants) is significantly associated with *PDX1* expression in bulk pancreas tissue according to GTEx v8 expression QTL (eQTL) results is consistent with it being a functional promoter variant (*P* = 2.45 x 10^-6^; **Figure 3A**).^32^ More specifically, the PDAC risk-increasing allele (A) was associated with decreased *PDX1* expression (**Figure 3A**). To test whether the PDAC GWAS and *PDX1* eQTL signals are mutually explained by the same underlying functional variant, we applied statistical colocalization analysis using the coloc R package^33^ that indicated both genetic association signals were very likely driven by the same functional variant (*PP*_H4_ = 0.976; **Figure 3C**). *PLUT* (also known as *PDX1-AS1*) is a non-coding antisense transcript whose expression tends to correlate with *PDX1* due to shared *cis*-regulatory elements and a direct role for the *PLUT* transcript in influencing local chromatin looping architecture to promote *PDX1* transcription.^34^ Notwithstanding this expected co-expression, rs9581943 was not significantly associated with *PLUT* expression in GTEx pancreas tissue (*P* = 0.0736) and the *PLUT* eQTL results were not well colocalized with the PDAC GWAS signal (*PP*_H4_ = 0.204; **Figures 3B** and **D**). However, it is worth noting that despite not reaching significance, the expression trend for *PLUT* does mirror that of *PDX1* with respect to rs9581943 genotype (**Figure 3B**).

**Figure 3:**
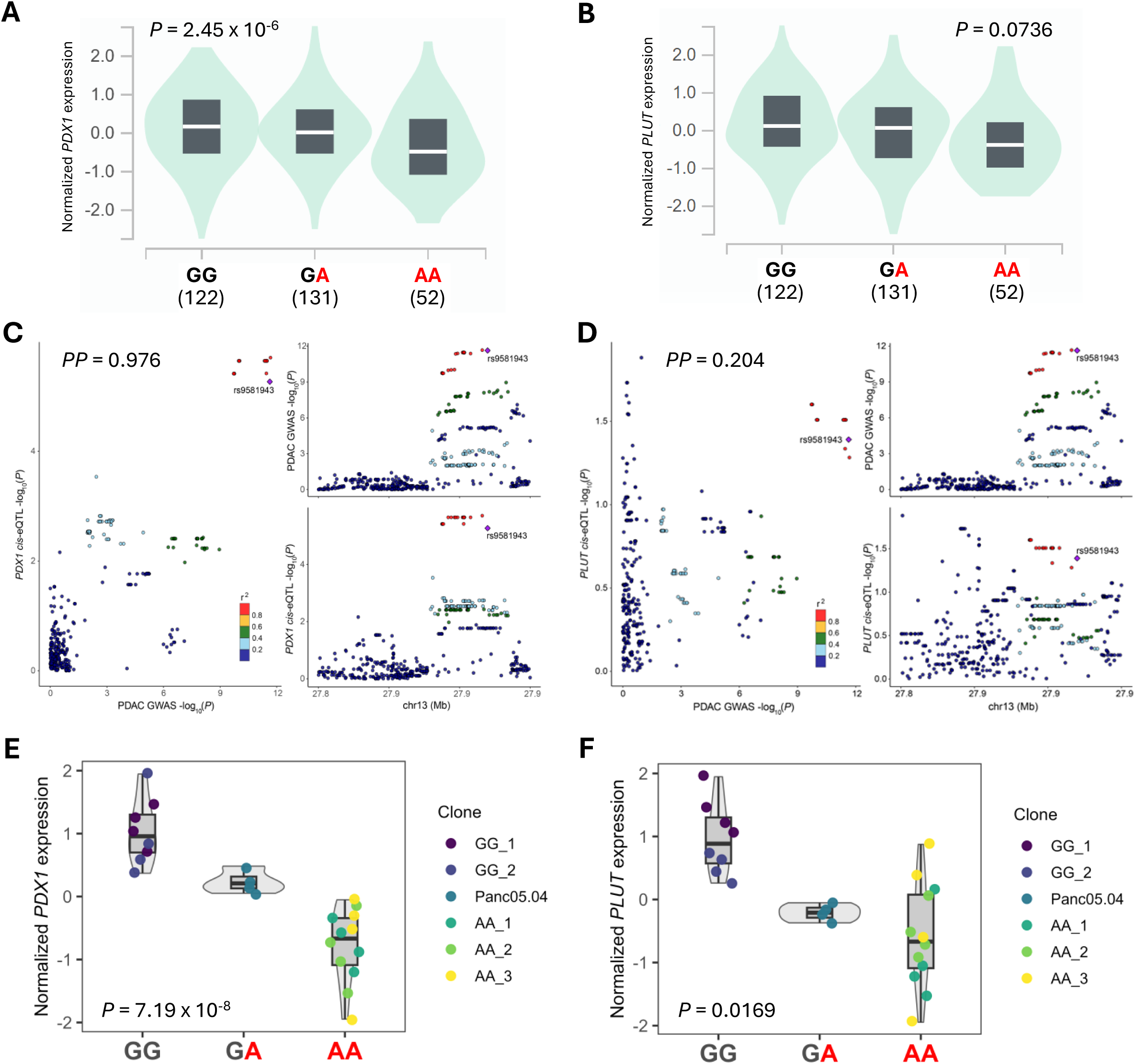
rs9581943 is functional *in situ* with transcriptional effects on *PDX1*. **(A-B)** Violin plots demonstrating GTEx v8 pancreas eQTL associations between rs9581943 genotype and *PDX1* **(A)** or *PLUT* **(B)** normalized expression. Numbers under each genotype indicate the sample count. **(C-D)** LocusCompare plots demonstrate the degree of colocalization between the PDAC GWAS signal and the *PDX1* **(C)** or *PLUT* **(D)** cis-eQTL signals (GTEx v8 pancreas). Posterior probability (*PP*) of a shared causal variant between signals was calculated with the coloc R package. SNPs are colored according to their r^2^ with rs9581943. **(E-F)** Violin plots demonstrating the associations between rs9581943 genotype in CRISPR/Cas9-edited Panc 05.04 clones and *PDX1* **(E)** or *PLUT* **(F)** normalized expression.

As with GWAS signals, LD among variants of an eQTL signal leaves underlying functional variant(s) uncertain, even with the implications of epigenetic regulatory marks and luciferase assay results. We therefore sought to directly test the causal relationship between rs9581943 and *PDX1* expression. CRISPR/Cas9 single-base editing was applied to the pancreatic cell line Panc 05.04 that is heterozygous for rs9581943 to yield clones homozygous for either its reference (G) or alternate (A) alleles. Expression of nearby genes were assayed by RT-qPCR and modeled by linear regression against all three genotypes (GG, GA, AA) in an otherwise isogenic background. The *PDX1* eQTL was significantly recapitulated among the edited clones with the PDAC risk increasing allele (A) again correlated with lower *PDX1* expression (*P* = 7.19 x 10^-8^; **Figure 3E**). *PLUT* expression was also significantly associated with rs9581943 genotype among these edited clones (*P* = 0.0169; **Figure 3F**), but the *CDX2* gene that lies ∼35kb downstream of *PDX1* did not indicate a significant correlation (*P* = 0.8534; **Figure S2**). We attempted to replicate these results in Capan-2, another pancreatic cell line heterozygous for rs9581943. The same trend was observed between the GA and AA genotypes, but we were unable to generate homozygous GG clones (**Figure S3**). Altogether, these results indicate rs9581943 genotype causally influences *PDX1* (and possibly *PLUT*) expression and, paired with the strong colocalization between GWAS and *PDX1* eQTL signals, suggest this causal influence likely mediates the PDAC risk effects at this locus.

### Overexpressing PDX1 in tumor-derived cell lines inhibits proliferation and increases apoptosis

To begin exploring the consequences of altered PDX1 expression in pancreatic contexts, we generated doxycycline-induced PDX1 overexpression clones from PANC-1 and MIA PaCa-2 cell lines (**Figure S4**). Cell proliferation was then measured for empty vector control and PDX1-inducible clones +/- doxycycline treatment. Overexpressing PDX1 led to notably decreased cell proliferation in both PANC-1 and MIA PaCa-2 backgrounds (**Figure S5**). Reduced proliferation could result from cell cycle blocks or increased cell death, so we examined cell cycle profiles and apoptosis frequencies in the control and PDX1-inducible cells +/- doxycycline treatment using flow cytometry. Cell cycle profiles indicated no notable changes in phase proportions between PDX1 overexpression induced and uninduced cultures (**Figure S6**). In contrast, the ratio of apoptotic to non-apoptotic cells increased significantly over time with PDX1 overexpression, suggesting the decreased cell proliferation was likely driven by increased apoptosis (**Figure S7**). While the results demonstrated that rs9581943 genotype modulates *PDX1* expression and that PDX1 overexpression constrains proliferation in tumor-derived cell lines, they do not reveal the cell-state contexts in which PDX1 exerts its endogenous functions. To map these contexts directly, we analyzed single-nucleus and single-cell RNA-seq datasets from normal, inflamed, and malignant pancreas.

### PDX1 buffers exocrine cell stress and fate in normal and inflamed pancreas

PDX1 is a key transcription factor required to establish the pancreatic progenitor domain during development and later acts as a master regulator of β-cell identity, insulin production, and glucose-responsive function.^35^ However, its role in terminally differentiated exocrine pancreas cells and functioning is less clear. As PDAC is believed to derive primarily from the exocrine compartment, we sought to clarify how altered *PDX1* expression might influence PDAC risk in acinar and ductal cells by analyzing the neonatal, adult, and chronic pancreatitis snRNA-seq pancreatic datasets reported by Tosti et al. (2021).^36^ After preprocessing the three datasets separately, exocrine cells were selected from each dataset and integrated into a shared embedding using FastMNN.^37^ Previously defined exocrine cell states were assigned based on unsupervised clustering and marker gene expression scoring (**Figure 4A** and **Table S4**). *PDX1* transcript detection was quite sparse and largely uninformative across these exocrine subtypes (**Figure 4A**), possibly due to infrequent transcriptional bursting combined with the typical sparsity of snRNA-seq, though PDX1 protein expression has been more consistently observable throughout the exocrine pancreas by immunohistological analyses.^38^ Consistent with protein-based observations, PDX1 expression regulatory activity levels inferred from expression of its target genes (see **Materials and Methods**) indicated continuous and structured PDX1 activity across the exocrine landscape (**Figure 4A**).

**Figure 4:**
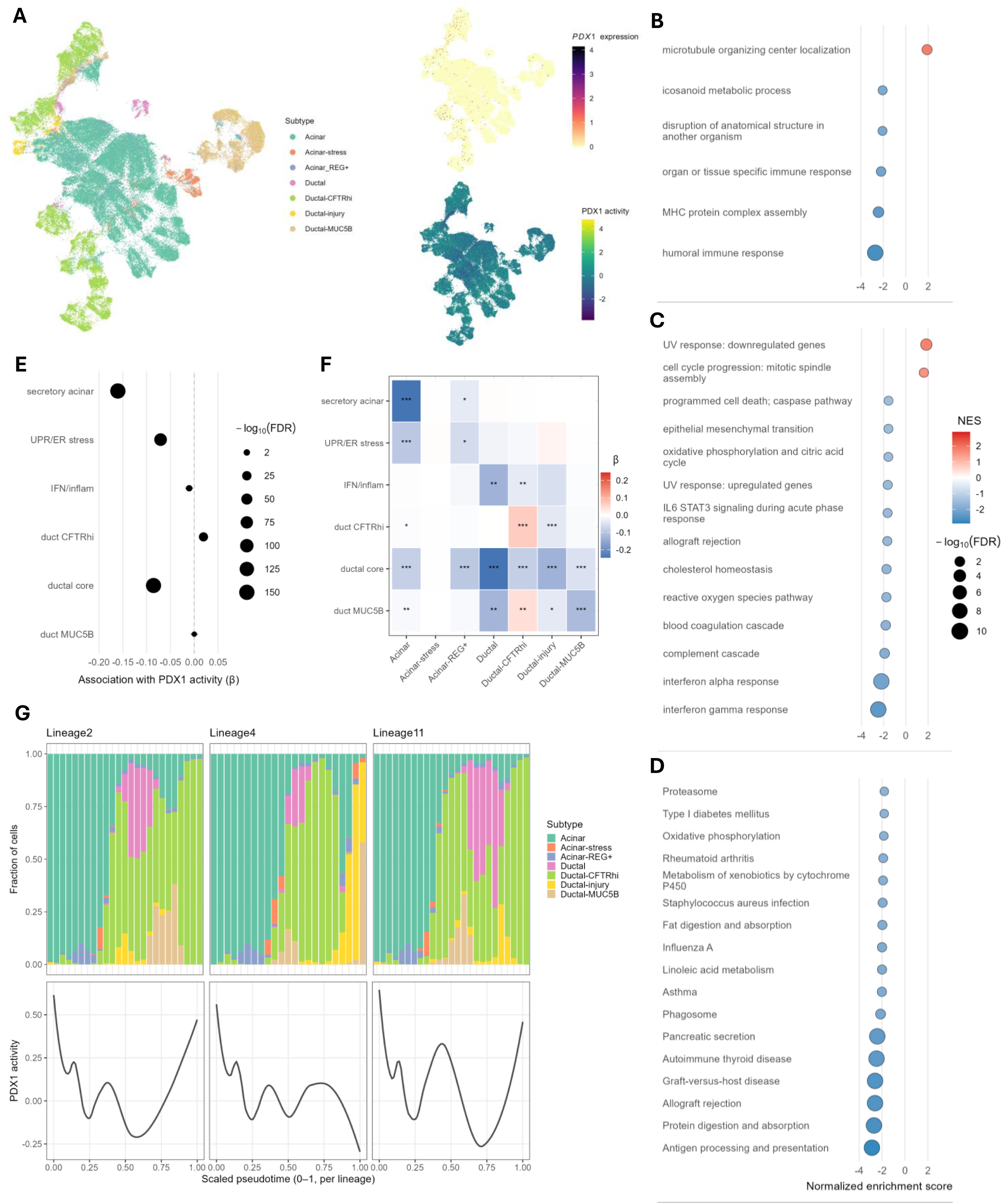
PDX1 activity associates with reduced exocrine cell stress and more homeostatic phenotypes. Key results from analyses of the Tosti et al. 2021 integrated neonatal, adult, and chronic pancreatitis snRNA-seq datasets. **(A)** UMAP plots of exocrine pancreatic cells colored by exocrine cell subtype, *PDX1* expression, or residualized PDX1 regulatory activity. **(B-D)** Forest plots of significantly enriched/depleted gene sets (FDR < 0.05) from the GO: Biological Processes **(B)**, MSigDB Hallmark **(C)**, and KEGG **(D)** gene set collections. Dot size scales with -log_10_(FDR) and dot shading corresponds to normalized enrichment score (NES). **(E)** Forest plot of global associations across all exocrine cells between residualized PDX1 activity and relevant gene module scores, where dot size scales with -log_10_(FDR). **(F)** Heatmap summarizing exocrine cell subtype-stratified associations between residualized PDX1 activity and relevant gene module scores. Grid shading indicates PDX1 activity coefficient effect size and *, **, and *** indicate FDR < 0.05, 0.01, and 0.001, respectively. **(G)** Stacked exocrine cell subtype composition plots over smoothed residualized PDX1 activity curves versus scaled pseudotime for each of three representative lineages inferred with the Slingshot R package.

Hurdle models were used to test association between residualized PDX1 activity (adjusted for confounding technical variables) and gene expression, and gene set enrichment analyses (GSEA) were performed against GO: Biological Processes^39,40^, MSigDB Hallmark^41^, and KEGG Pathways^42^ gene set collections to investigate biological consequences of varying PDX1 activity (**Figures 4B-D** and **Tables S5-S8**). This suggested PDX1 activity associated with depletion of secretory enzyme production, mitochondrial respiration, inflammatory signaling, and epithelial to mesenchymal transition (EMT) gene sets which prompted investigation of PDX1 activity’s association with gene modules representing a high acinar secretory load (secretory acinar), unfolded protein response/endoplasmic reticulum stress (UPR/ER stress), interferon/inflammatory signaling (IFN/inflam), a core ductal program (ductal core), a high CFTR ductal state (duct CFTRhi), and a mucinous ductal state (duct MUC5B) (**Table S9**). Consistent with the GSEA results, across all cells globally we observed significant downregulation of the secretory acinar, ductal core, UPR/ER stress, and IFN/inflam gene modules, but significant upregulation of the duct CFTRhi gene module (**Figure 4E**). Despite the positive association with ductal modules and negative association with the secretory acinar module, these results do not imply PDX1 activity encourages acinar-to-ductal metaplasia (ADM) because the regression models were adjusted for exocrine cell subtype, indicating these associations hold even when subtype is held constant. We therefore stratified the gene module-to-PDX1 activity modeling by subtype to resolve PDX1’s putative influence within these distinct exocrine cell states (**Figure 4F**). Within acinar and acinar-REG+ cells, PDX1 activity was associated with reduced secretory and UPR/ER stress programs, and importantly, this occurred without concurrent induction of ductal identity programs. The story is a little more subtle among ductal subtypes.

The ductal core module represents a KRT+/MUC1+/EPCAM+ simple epithelial program that is shared across both classical intralobular and early metaplastic ducts (**Table S9**).^36,43,44^ Therefore, the negative association with the ductal core and IFN/inflam modules together with the positive association with the duct CFTRhi module within the ductal-CFTRhi subtype is consistent with PDX1 helping to maintain a healthier homeostatic ductal cell state (**Figure 4F**). Indeed, PDX1 activity was negatively associated with the ductal core module across all ductal subtypes, emphasizing that PDX1 is not primarily pushing cells towards the simple epithelial duct program but seemingly favors the healthier ductal-CFTRhi cell state (**Figure 4F**). Strikingly, PDX1 activity was negatively associated with the duct MUC5B module across most ductal subtypes, including the ductal-MUC5B subtype itself, but was positively associated with the duct MUC5B module in ductal-CFTRhi cells (**Figure 4F**). This apparent contradiction may be resolved if ductal-CFTRhi cells with higher duct MUC5B module scores represent a transitional cell state wherein increased PDX1 activity may function reactively to push ducts towards the more adaptive, homeostatic ductal cell state with lower metaplastic potential. To better test this hypothesis, we performed RNA trajectory and pseudotime inferences from acinar to ductal cell states using the Slingshot R package and identified three main representative lineages. By comparing the PDX1 activities to exocrine subtype proportions as pseudotime increases along these lineages, across all three lineages we observed PDX1 activity was high in stable states (acinar, ductal-CFTRhi), fell during transitions into stress or injury programs, and then increased again as CFTRhi recovery phases emerged (**Figure 4G**). Taken together, these results suggest PDX1 activity may provide a buffering mechanism to the exocrine pancreas, alleviating high secretory load/ER-stressed states in acinar and biasing ductal cells toward less inflammatory, more homeostatic CFTRhi phenotypes as opposed to mucinous or injury-related programs.

### PDX1 promotes more epithelialized Classical pancreatic tumor cell states

To test PDX1’s influence on cell states among transformed exocrine-derived epithelial cells, we analyzed the pancreatic cancer scRNA-seq dataset produced and reported by Chan-Seng-Yue et al. (2020).^45^ After preprocessing and RPCA integration of the 15 samples into a shared embedding, tumor cells were selected and typed as Classical, Hybrid, and Basal using published NMF-signatures (**Figure 5A** and **Table S10**).^45^ This 3-family subtyping scheme was more robust and interpretable for our purposes while preserving the key transcriptional continua (from Classical to Basal) observed in all tumors tested. As for the Tosti et al. datasets, *PDX1* transcript detection was quite sparse but inferred PDX1 activity was continuous and structured across all cells (**Figure 5A**). Stratifying PDX1 activity by tumor subtype demonstrated a clear trend with higher activity in Classical cells, lower in Basal, and Hybrid cells in between, suggesting PDX1 activity associates with classical epithelial maintenance but is diminished along more aggressive, basal-like trajectories (**Figure 5B**). To better resolve this at a pathway level, we again used hurdle models to test the association between PDX1 activity and gene expression and performed GSEA to identify up- or down-regulated gene sets among the GO: Biological Processes^39,40^, MSigDB Hallmark^41^, and KEGG^42^ pathways collections (**Figures 5C-E** and **Tables S11-S14**). Consistent with our observations in normal exocrine pancreas cells, we detected negative associations with ribosomes, protein translation, mitochondrial respiration, inflammation/immune activation, and epithelial stress pathways, suggesting PDX1 activity may lower metabolic strain and reduce inflammation (**Figures 5C-E** and **Tables S12-S14**). Furthermore, PDX1 activity was positively associated with epithelial differentiation and adhesion programs, consistent with PDX1 pushing cells towards a less invasive and more epithelialized classical tumor cell state (**Figures 5C-E** and **Tables S12-S14**). Note that, though PDX1 activity was positively associated with the “epithelial mesenchymal transition” gene set as defined by MSigDB, the genes driving the enrichment consisted largely of integrins, laminins, and collagens rather than traditional mesenchymal markers, and is therefore consistent with PDX1 biasing cells towards an epithelial phenotype (**Table S15**). Similarly, the genes driving the enrichment of the “cell cycle progression: mitotic spindle assembly” gene set consisted of mainly microtubule/centrosome and polarity-related genes and is therefore consistent with PDX1 activity’s negative association with the Hallmark “cell cycle progression: E2F targets” gene set that implies reduced proliferative drive (**Table S15**). Altogether, these results suggest that loss of PDX1 activity may facilitate de-differentiation, invasiveness, and inflammation, leading to more aggressive pancreatic tumor cell states.

**Figure 5:**
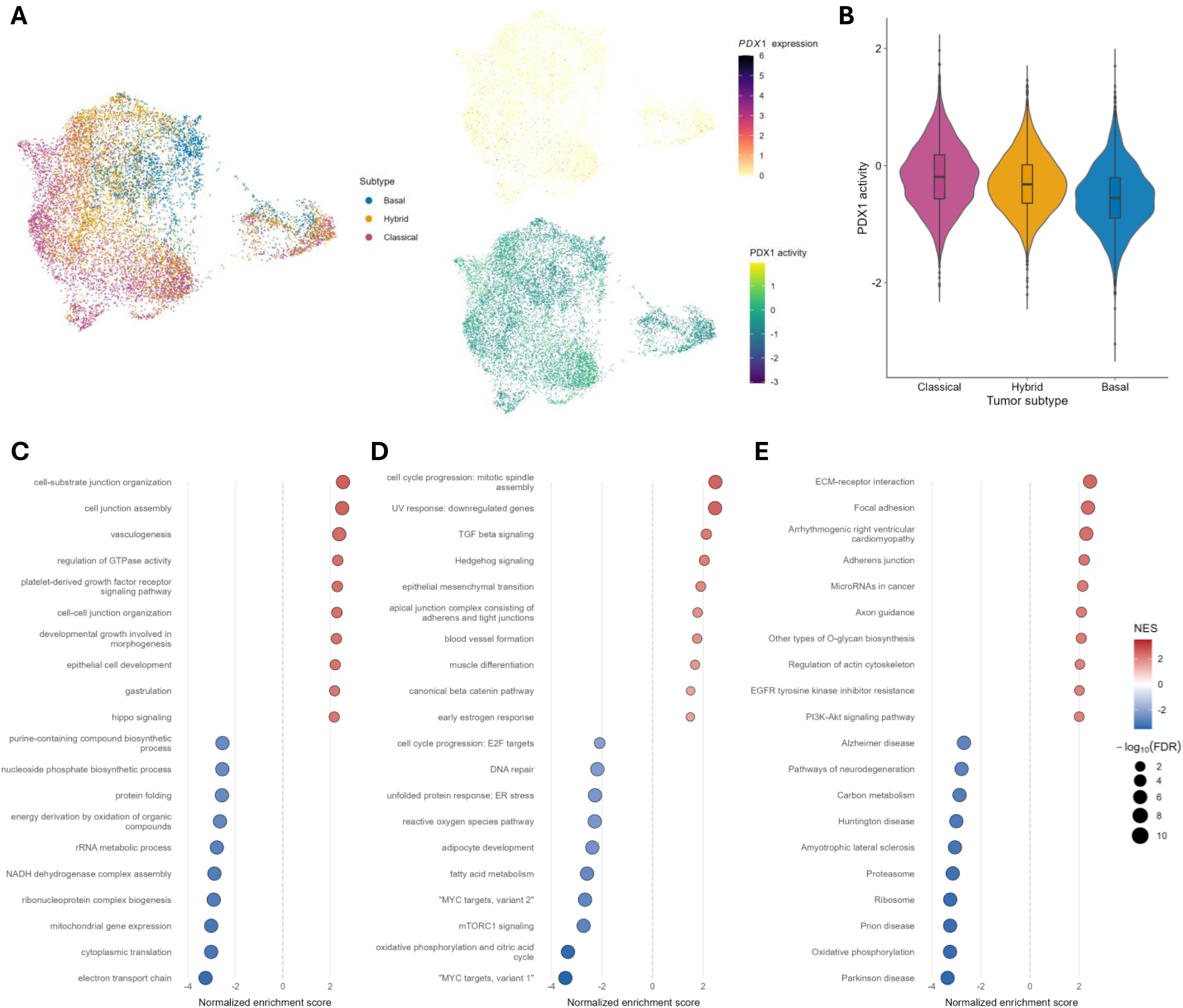
PDX1 activity associates with less aggressive and more epithelialized phenotypes in pancreatic tumor cells. Key results from analyses of the Chan-Seng-Yue et al. 2020 integrated pancreatic tumors scRNA-seq dataset. **(A)** UMAP plots of pancreatic tumors cells colored by pancreatic tumor cell transcriptional subtype, *PDX1* expression, or PDX1 regulatory activity. **(B)** Violin plot of PDX1 regulatory activity stratified by pancreatic tumor cell transcriptional subtype. **(C-E)** Forest plots of top 10 significantly (FDR < 0.05) enriched or depleted gene sets from the GO: Biological Processes **(C)**, MSigDB Hallmark **(D)**, and KEGG **(E)** gene set collections. Dot size scales with-log_10_(FDR) and dot shading corresponds to normalized enrichment score (NES).

## Discussion

Our study has implicated rs9581943 as a likely functional driver at the chr13q12.2 *PLUT*/*PDX1* pancreatic cancer GWAS signal. The rs9581943 risk-increasing allele reduces *PDX1* expression, demonstrated experimentally by single-base CRISPR/Cas9 editing of pancreatic cell lines. Investigation of its role in exocrine pancreas cell contexts using snRNA-seq datasets suggested PDX1 activity buffers against stressed states and stabilizes homeostatic phenotypes in both acinar and ductal cells, while in pancreatic tumors PDX1 activity correlated with less inflamed and more epithelialized tumor cell states. Altogether, we propose an explanatory model for this PDAC GWAS signal in which the risk-increasing allele of rs9581943 diminishes the transcriptional responsiveness of the *PDX1* gene, consequently eroding the capacity for PDX1 to buffer against stressed exocrine cell states with more metaplastic potential, which over time increases risk of malignant transformations.

This aligns well with current injury-driven models for PDAC evolution that suggest chronic ER stress in acinar cells or other sources of inflammation can trigger acinar-to-ductal metaplasia (ADM) that may progress to PanIN formation and ultimately lead to malignant PDAC transformation.^46^ PDX1’s role in that injury-driven trajectory has been considered previously, and positive correlation between *PDX1* expression and ADM has been reported.^47,48^ Given PDX1’s well established roles in early pancreas formation and pancreatic progenitor cell specification, it has been suggested that *PDX1* expression in stressed acinar cells may induce a more progenitor-like state leading to ADM.^35,48,49^ That interpretation conflicts with our model of PDX1 activity buffering against stress and stabilizing homeostatic exocrine cell states, but such uncertainty about whether PDX1 is tumor suppressive or oncogenic is not new. In addition to experiments linking PDX1 expression to ADM and PanIN formation, it was demonstrated that PDX1 is required for survival of established PDAC tumors, suggesting an oncogenic influence.^50^ However, the same study also observed that conditional acinar-specific *Pdx1* deletion in adult transgenic mice followed by cerulein induction of pancreatitis accelerated ADM and PanIN formation. Furthermore, while control mice were able to regenerate their acinar after recovery from cerulein injury, the *Pdx1* deleted mice did not. Therefore, Roy *et al*. concluded that PDX1 is important for acinar maintenance and regeneration after ADM, which nicely accords with our proposed model for PDX1 in exocrine pancreatic cells. An earlier biochemical study has also shown that PDX1 functions in pancreatic acinar cells through highly cooperative transcriptional complexes, in which transcriptional output is an order of magnitude greater than that of PDX1 alone and is exquisitely sensitive to PDX1 dosage and activation potential.^51^ This implies that even modest differences in the transcriptional responsiveness of the *PDX1* gene, as observed across rs9581943 genotypes, could have a disproportionate effect on acinar transcriptional programs.

If PDX1 is important for acinar maintenance, as suggested by our study and that of Roy *et al*. (2016)^50^, then the observed increased PDX1 expression in ADM lesions might represent a reactive attempt to re-establish a homeostatic acinar state. This would be consistent with our lineage analyses that indicated PDX1 activity starts rising before injured/stress acinar or ductal populations begin diminishing (**Figure 4G**). Similarly, our proposed explanation for the positive correlation with the duct MUC5B gene module only observed in Ductal-CFTRhi cell population is that ductal cells with expression of both CFTRhi and MUC5B gene modules represent a transitional fringe in which PDX1 activity is specifically increased to drive cells toward the more homeostatic Ductal-CFTRhi state. This hypothesis is supported by a recent study using a cystic fibrosis ferret model that revealed CFTR loss triggers increased PDX1 expression in ductal epithelium, suggesting the presence of a feedback mechanism between CFTR and PDX1 in ductal cells.^52^

It may not be surprising that PDX1’s role in adult exocrine cell states has remained elusive. Even as single cell methodologies have enabled unprecedented cell state resolution, the sparsity of detected *PDX1* transcripts in acinar and ductal cells has made it technically challenging to characterize its exocrine roles. In such scenarios, expression metrics tend to be very poor proxies of TF activity. However, such challenges are surmountable by activity inferences that are demonstrably more robust, which follows from inferred activities being derived from the coordinated expression of dozens to hundreds of target genes.^53,54^ The ability to recover coherent and meaningful structure for PDX1 activity across cell states was key to revealing new insights into PDX1’s exocrine roles. Conversely, the emergence of a coherent and biologically sensible PDX1 activity landscape across datasets from neonatal, adult, inflamed, and malignant tissues strengthens the argument that these inferred activities reflect genuine regulatory programs rather than analytical artifacts. Taking similar activity-based approaches may prove critical when investigating other lowly expressed TFs moving forward.

As remarked in the Introduction, one of the clearest patterns observed among several PDAC GWAS signals published to date is their proximity to TF genes important for pancreas development and homeostasis, such as *NR5A2*, *HNF1A*, *HNF1B*, *HNF4G*, and *PDX1*.^7,12,19–24^ Like *PDX1*, some of these have been identified as marker genes for more epithelialized and less aggressive Classical or Pancreatic Progenitor-like tumor subtypes.^26,27,36^ Going forward, it would be interesting to explore whether those TFs also play complimentary or cooperative roles in the exocrine pancreas to buffer against stress and stabilize homeostatic epithelial phenotypes. However, unlike *PDX1*, those other pancreatic TF genes do not have significant *cis*-eQTLs that overlap, let alone colocalize, with their nearby PDAC GWAS signals, necessitating alternative approaches (e.g., activity QTL analysis) to sufficiently characterize their signals.^55,56^

Interestingly, this chr13q12.2 PDAC GWAS signal appears to influence the UPR/ER stress pathway via PDX1. Similarly, two other PDAC GWAS signals extensively characterized in published post-GWAS functional studies (chr16q23.1 and chr1p36.33) were shown to influence protein accumulation and inflammatory signaling pathways, which are coupled by UPR/ER stress pathways.^14,17^ The UPR/ER stress response therefore seems to be emerging as a common pathway for mediating the genetic component of PDAC risk.

Several questions relevant to this study remain open for future investigations. First, we were unable to identify TF(s) responsible for the rs9581943 allele-preferential expression regulatory effects. *In silico* TF binding predictions yielded many potential allele-preferential TFs, but none could be convincingly validated (data not shown). This difficulty may be explained by recent work indicating functional variants often alter non-canonical TF binding motifs flanking more canonical motifs wherein the non-canonical motifs serve to widen the attractor region for a given TF.^57^ Second, the luciferase reporter assays indicated the rs9581943 risk allele yielded higher luciferase expression, which was opposite to the direction of effect observed for *PDX1* expression in our GTEx eQTL and CRISPR/Cas9 analyses. This may be because these luciferase reporter assays use synthetic plasmid constructs that are episomal and lack chromatin structures while the eQTL and CRISPR/Cas9 experiments better reflect the endogenous context. Third, while evidence for rs9581943’s functional relevance for this GWAS signal is strong, we cannot exclude the possibility that other variants in the signal’s credible set may also serve some relevant functions in certain contexts. Similarly, we cannot exclude *PLUT* as a potential target gene for this GWAS signal, though based on the literature it would probably function by modulating *PDX1* expression as well.^34^ Finally, even though PDAC is believed to primarily derive from the exocrine pancreatic compartment, it is possible some component of this GWAS signal’s PDAC risk effects may be mediated indirectly by modulating PDX1 activity in β-cells since insulin/IGF signaling is known to influence tumor biology.^58,59^

## Materials and Methods

### Statistical fine-mapping of the chr13q12.2 PDAC GWAS locus

The Sum of Single Effects (SuSiE)^28,29^, a Bayesian approach, was used on the PanScan I+II, PanScan III, and PanC4 meta-analyzed GWAS summary statistics to identify credible sets of variants likely harboring functional variant(s). Using a threshold of 0.95 probability that the credible set of variants contains a causal SNP, SuSiE models for L = 1, 2, or 3 expected credible sets were tested. Because setting L = 2 or 3 forced SuSiE to allocate multiple components within a single, tightly linked association peak (clear over-splitting), we fixed L = 1 to model the locus as a single causal signal, which is consistent with the LD structure at this region. PanScan I+II data^3^ (including cases and controls) were used to generate the required LD matrix using PLINK v1.9.^60^

### Cell lines and engineered clones

Four pancreatic tumor-derived cell lines (PANC-1, MIA PaCa-2, Panc 05.04, and Capan-2) along with HEK293T and HeLa cells were purchased from ATCC (Gaithersburg, MD, USA) and cultured as recommended (www.atcc.org).

MIA PaCa-2 and PANC-1 cell lines were stably transfected with a TetOn3G transactivator (pLVX-Tet3G; Takara Bio USA, San Jose, CA, USA) via lentiviral transduction (MOI = 3) and selected with 5µg/ml blasticidin. Human *PDX1* cDNA was sub-cloned into the lentiviral Tet response vector (pLVX-TRE3G; Takara Bio USA). The resulting construct-containing or empty vectors were stably transduced into the TetOn3G expressing cells (MOI = 1) and selected with 2µg/ml puromycin. Clones were isolated, expanded and tested for PDX1 inducibility by Western analysis.

For CRISPR/Cas9 single-base editing, Cas9-sgRNA ribonucleoprotein (RNP) complex was prepared by mixing recombinant Cas9 protein with IVT-1686 sgRNA (**Table S16**) at final concentrations of 6.7 µM and 20 µM, respectively, in 1X TE buffer. Panc 05.04 and Capan-2 cell lines were cultured to 80% confluency and transfected with 3 µL of Cas9-sgRNA RNP complex and 1 µg of donor oligonucleotide (Oligo-2547 for G->A, or Oligo-2548 for A->G; **Table S16**) by electroporation using the Lonza 4D-Nucleofector X Unit with the SE Cell Line 4D-Nucleofector X Kit S (Lonza, Basel, Switzerland). After 2 days of recovery, clonal populations were isolated by limiting dilution in 96-well plates and expanded. Clones were genotyped by qPCR with TaqMan (ThermoFisher Scientific, Rockville, MD, USA) rs9581943 genotyping assay (C 27845322_20; **Figure S1**).

### Luciferase reporter cloning and assays

DNA fragments 61bp long and centered on rs2297316, rs9579218, and rs9581943 were prepared by annealing complimentary oligonucleotides (Integrated DNA Technologies, Coralville, IA, USA) (**Table S16**). The DNA fragments containing the cluster of seven SNPs (rs7993114, rs11618581, rs9554197, rs11618036, rs9579127, rs7999100, rs8000004), were ordered as premade, double-stranded synthetic genes (Integrated DNA Technologies) (**Table S16**). Inserts were cloned into the KpnI and NheI sites of the pGL4.23[luc2/minP] (Promega, Madison, WI, USA) luciferase vector in the 5’-to-3’ or 3’-to-5’ orientation. All plasmid inserts were sequence-verified to contain the correct inserts and genotypes. The Firefly reporter plasmids (and a Renilla luciferase control vector) were co-transfected into PANC-1 and MIA PaCa-2 pancreatic cancer cell lines at ∼40% confluence using Lipofectamine 3000 (ThermoFisher Scientific). Luciferase activity was measured 48hrs after transfection with the Dual Luciferase Reporter Assay System (Promega). Firefly luciferase activity was normalized to the Renilla luciferase activity. Each experiment was performed in 6-8 replicate wells and repeated at least three times. A t-test was performed to assess the significance for allelic differences in luciferase activity.

### Electrophoretic mobility shift assays (EMSA)

SNP-centered 31 bp oligonucleotides (**Table S16**) were labeled with IRDye©700 fluorescent dye on the 5’ end and HPLC purified (Integrated DNA Technologies). Competitor oligonucleotides (**Table S16**) were unlabeled (Integrated DNA Technologies). Oligonucleotides were annealed at 99 °C and cooled slowly to room temperature. EMSA binding reactions were as follows: 10X binding buffer, 50% glycerol, polyDiDC, nuclear lysate (2.5–5 μg, ActiveMotif, Carlsbad, CA, USA), labeled oligo (5 nM), water. Competition binding reactions included unlabeled oligos at 50X and 100X of the labeled oligo concentration. Reactions were incubated at room temperature in the dark for 20 min. Reactions were then loaded on 4–12% gradient TBE gels (ThermoFisher Scientific) with 0.5X TBE and ran for 100 min at 90 V. Gels were imaged on the BioRad ChemiDoc^TM^ (BioRad, Hercules, CA, USA) with the IR680 setting.

### Colocalization analysis between GWAS and eQTL signals

Colocalization analysis was performed on the 2018 GWAS summary statistics^7^ and the GTEx v8 pancreas eQTL data (downloaded from the GTEx portal)^32^ using the coloc R package (version 4.0).^33^ All SNPs from chr13:27,800,000-27,900,000 (hg38) were used after harmonizing the GWAS and eQTL results.

### RNA extraction and RT-qPCR gene expression analysis

RNA was isolated using the QIAGEN RNeasy kit with a DNase digest and the QIAcube (QIAGEN, Germantown, MD, USA). RNA was reverse transcribed to cDNA using SuperScript III Reverse Transcriptase (Thermo Fisher Scientific). Gene expression levels were quantified by qRT-PCR using TaqMan (Thermo Fisher Scientific) assays: *PDX1* (Hs00236830_m1), *PLUT* (Hs04940031_m1), *CDX2* (Hs01078080_m1), *HPRT* (Hs99999909_m1).

For each gene, ΔC_t_ expression values (normalized to *HPRT1*) were rank-based inverse normal transformed (INT) across all samples. Associations between rs9581943 genotype (modeled additively as A allele dosage) and transformed expression in CRISPR/Cas9-edited Panc 05.04 clones were tested using linear mixed-effects models of the form:

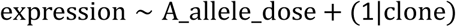

### Cell proliferation assays

PDX1-inducible and uninducible control clones were plated in 12-well plates 24 hours prior to the addition of 100 ng/mL doxycycline (Takara Bio USA) to the +Dox cultures, which marked the beginning of the time course. Cell images were taken of each well using the Agilent BioTek Lionheart FX plate reader (Agilent, Santa Clara, CA, USA) twice a day for 5 or 6 days. Cells were counted and cell growth curves were plotted with the accompanying software (Agilent).

### Cell cycle assays

Cultures of PDX1 inducible or uninducible control clones were grown to ≤90% confluency for 0–3 days with 0 or 100 ng/mL doxycycline (Takara Bio USA). Cells were trypsinized, washed in 1x phosphate-buffered saline (PBS) and fixed in 70% ethanol in 1x PBS at −20°C overnight. After washing twice in 1x PBS, cells were resuspended in 0.5 or 1.0 mL 1x PBS. RNase A (ThermoFisher Scientific) was added to 1 μg/mL and cells incubated for 1 h at 37°C, after which propidium iodide (Sigma-Aldrich, Saint Louis, MO, USA) was added to 40 μg/mL. At least 30,000 cells for each sample were counted and measured for propidium iodide fluorescence on an Attune NxT flow cytometer (ThermoFisher Scientific). Cell cycle phase populations were estimated after doublet discrimination using ModFit LT (http://www.vsh.com/products/mflt/).

### Apoptosis assays

Apoptosis assays were performed across three independent experiments using the Dead Cell Apoptosis Kit with Annexin V for Flow Cytometry (ThermoFisher Scientific) run on an Attune NxT Flow Cytometer (ThermoFisher Scientific). PDX1-inducible and control clones of PANC-1 and MIA PaCa-2 were grown +/-doxycycline for 24 hours prior to harvesting, staining, and flow cytometry. Apoptosis rates were reported as the ratio of apoptotic cells (positive for both Alexa Fluor 488 and Propidium Iodide) to normal live cells (negative for both Alexa Fluor 488 and Propidium Iodide).

### snRNA-seq analysis (Tosti et al. 2021 datasets)

We reanalyzed the neonatal, adult, and chronic pancreatitis (CP) human pancreas single-nucleus RNA-seq datasets reported by Tosti et al. (2021).^36^ All analyses were performed in R (v4.5.1) using the Script_for_Tosti_snRNA-seq_analyses_used_in_Hoskins_et_al_2026.R available in this study’s GitHub repository (https://github.com/hoskinsjw/PDX1_post-GWAS_study). Details may be found in the script file, but brief summaries of key processing steps and analyses are provided below.

#### Preprocessing, RNA contamination estimation, and quality control

For each of the three contexts (neonatal, adult, and chronic pancreatitis (CP)), we started from the published Seurat objects and processed samples with a common pipeline. Briefly, we first sanitized cell names to ensure uniqueness and alignment between expression matrices and metadata, then computed per-cell QC metrics including the percentage of mitochondrial and ribosomal transcripts. Ambient RNA contamination was estimated per sample using DecontX (celda)^61^, with sample identity and an initial clustering used as covariates.

We then applied a unified, adaptive cell-level filtering step that applies sample-wise median-absolute-deviation (MAD) based trimming on the number of detected genes, and enforces upper bounds on mitochondrial and ribosomal percentages, and on DecontX contamination. In our final analysis we used 3 MADs, a minimum of 150 detected genes per nucleus, and upper thresholds of 10% mitochondrial, 35% ribosomal RNA, and 50% DecontX contamination.

#### Context-specific integration and metadata harmonization

Within each context (neonatal, adult, CP), we integrated nuclei across samples using batchelor’s fastMNN implementation.^37^ Highly variable genes (n = 3,000 per context) were selected, and 30 principal components (PCs) were computed from the MNN-corrected expression followed by a 2D UMAP embedding (30 PCs, 30 neighbors, minimum distance 0.3). We then constructed a shared-nearest-neighbor graph on the MNN PCs and performed unsupervised clustering at resolution 0.5.

To make downstream integration across contexts easier, we harmonized sample-level metadata (dataset label, context, donor, sample ID, and anatomical region) using a dedicated harmonize_metadata helper, which maps Tosti’s original annotations into a consistent set of columns (dataset, context, donor, sample, anatomical_region, and celltype_curated).

#### Exocrine cell selection and combined integration

We next restricted the analysis to exocrine cells (acinar and ductal) using both the curated Tosti cell types and marker-based module scoring. Briefly, we defined panels of acinar and ductal marker genes largely based on Tosti et al. (2021)^36^ and implemented an exocrine_subset helper that selects clusters enriched for these modules while excluding endocrine, stromal, and immune lineages. The resulting exocrine subsets from neonatal, adult, and CP contexts were then merged at the counts level and re-integrated using fastMNN (batch = sample_ID) to generate a joint exocrine PCA and UMAP (**Figure S8**). We re-clustered the integrated exocrine object at resolution 0.5 for use in subsequent subtype assignment and trajectory analyses.

#### Exocrine subtype assignment and axis scoring

Exocrine subtypes were assigned using a marker-panel scoring approach. We defined eight axes corresponding to transcriptionally coherent exocrine programs defined in **Table S4**. For each axis, we computed a Seurat module score (log-normalized RNA assay), tracked the resulting metadata columns, and assembled a cell-by-axis matrix of scores. Per cell, we assigned the axis with the maximum score and defined a confidence metric as the margin between the top two axes. This axis was then mapped to a human-readable subtype label (Acinar, Acinar_REG+, Acinar-stress, Ductal, Ductal-CFTRhi, Ductal-MUC5B, Ductal-injury) and a discrete confidence category (very_low, low, moderate, high) based on the score margin. The final subtype label and axis score matrix were saved for downstream embeddings and module analyses (**Figure S8B**). Exocrine cell subtype proportions within each context (i.e., neonatal, adult, and CP) are shown in **Figure S9**.

#### Tuned combined embedding and WNN graph

To obtain an embedding that simultaneously preserves global structure and the exocrine subtype axes, we combined two low-dimensional representations: MNN PCs and axis-based PCs. We first computed a PCA on the axes score matrix and a UMAP from these axes PCs. We then used a tuning procedure to generate a combined PC space and UMAP that interpolate between the MNN-based and axis-based representations, selecting weights that optimize a balance between subtype separation and context mixing. UMAPs inferred from the MNN-based, subtype axis-based, and tuned combined PC spaces demonstrate donor-level integration and subtype clustering produced by each embedding (**Figure S10**). The tuned combined embedding was used for visualization in **Figure 4A** and downstream trajectory and pseudotime inferences. In parallel, we built a weighted nearest-neighbor (WNN) graph combining MNN and axes PCs using Seurat’s FindMultiModalNeighbors and FindClusters on the WNN graph; the WNN UMAP served as a diagnostic check that the tuned embedding remained concordant with the graph structure.

#### PDX1 expression regulatory activity inference and residualization

Because *PDX1* transcripts are sparse in snRNA-seq, we inferred PDX1 expression regulatory activity from its target genes. We constructed a union regulon collection by combining human DoRothEA (confidence levels A–C)^62^ and CollecTRI^63^ TF–target annotations, then ran VIPER^53,54^ to compute TF activity scores for all TFs.

To reduce technical confounding, we residualized all TF activity z-scores for five covariates: number of detected genes per cell (cngeneson), percent mitochondrial RNA, percent ribosomal RNA, DecontX contamination, and PC1 from the integrated MNN PCA. For each TF we fit a linear model of the form:

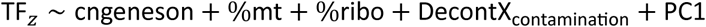

and stored the residuals. The residualized PDX1 activity was used as our primary measure of PDX1 regulatory activity in all downstream association analyses and visualizations. Diagnostics confirmed that this residualization substantially attenuated correlations between PDX1 activity and technical covariates while preserving biological variation across subtype, context, and donor.

#### Hurdle models and gene set enrichment analysis (GSEA)

We used a two-part hurdle framework to quantify associations between residualized PDX1 activity and per-gene expression across exocrine cells. For each gene, we defined:

- a binary detection indicator (expressed vs. not expressed),
- log-normalized expression among detected cells.

Using the log-normalized RNA matrix and aligned metadata, we fit for each gene a logistic model for detection and a linear model for expression among positives:

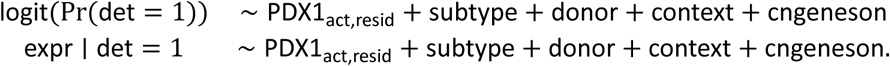

Models were fit in parallel across genes with a minimum of 25 expressing cells per gene. We extracted the PDX1 coefficient from each component, obtaining a slope and *P*-value for detection (log-odds) and continuous expression. Detection and continuous statistics were then combined with equal weighting into signed z-scores and an overall *P*-value was calculated by Fisher-style combination (**Table S5**).

For GSEA, we exported ranked gene lists based on the signed combined z-scores and ran pre-ranked GSEA^64^ run externally with WebGestalt (https://www.webgestalt.org/#) against GO: Biological Processes^39,40^, MSigDB Hallmark^41^, and KEGG^42^ gene set collections. Enrichment results were aggregated into collection-specific tables and plotted as dotplots (**Figures 4B–D** and **Tables S6–S8**).

#### Gene modules and module–PDX1 association models

To summarize key biological programs highlighted by GSEA, we defined curated gene modules representing high acinar secretory load (secretory acinar), unfolded protein response / ER stress (UPR/ER stress), interferon/inflammatory signalling (IFN/inflam), a shared ductal core program, a CFTR-high ductal state (duct CFTRhi), and a mucinous MUC5B-high ductal state (duct MUC5B), as described in **Table S9**. Module scores were computed using AddModuleScore. For global associations (**Figure 4E**), we regressed each module on residualized PDX1 activity and technical covariates:

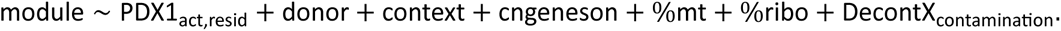

For subtype-stratified analyses (**Figure 4F**), we fit analogous models separately within each exocrine subtype.

#### Trajectory inference and pseudotime

To evaluate how PDX1 activity varies along acinar–ductal transitions, we performed trajectory inference with the slingshot R package.^65^ Starting from the integrated exocrine Seurat object, we converted counts and metadata to a SingleCellExperiment, carrying over the tuned combined PCA and UMAP embeddings. We then ran slingshot on the combined PCs using the WNN/Seurat cluster labels and automatically selected a root cluster enriched for acinar cells (highest fraction of Acinar and Acinar_REG+ subtypes) (**Figure S11**). Pseudotime curves were computed across all Slingshot lineages and stored in cell × lineage matrices.

To prioritize biologically plausible acinar-to-ductal trajectories, we computed lineage-level metrics including the number of cells per lineage, pseudotime range, and change in ductal-like subtype fraction from early to late pseudotime, and we down-weighted highly correlated, redundant lineages using a lineage–lineage pseudotime correlation matrix. A wrapper function combined these metrics to select three distinct representative lineages (Lineages 2, 4, and 11) that spanned long pseudotime ranges and exhibited increasing ductal-like composition. These lineages were used to generate the PDX1 activity and subtype composition trends along pseudotime shown in **Figure 4G**.

### scRNA-seq analysis (Chan-Seng-Yue et al. 2020 dataset)

All tumor scRNA-seq analyses were performed in R (v4.5.1) using the Script_for_Chan-Seng-Yue_scRNA-seq_analyses_used_in_Hoskins_et_al_2026.R available in this study’s GitHub repository (https://github.com/hoskinsjw/PDX1_post-GWAS_study). Details may be found in the script file, but brief summaries of key processing steps and analyses are provided below.

#### Preprocessing, quality control, DecontX, and integration

Raw 10x Genomics HDF5 count matrices for the 15 primary pancreatic tumors profiled by Chan-Seng-Yue et al. (2020)^45^ were imported into Seurat as separate objects. For each sample, per-cell quality-control metrics were computed, including the percentage of reads mapping to mitochondrial and ribosomal genes. Cells with <500 detected genes, ≥25% mitochondrial RNA, or ≥20% ribosomal RNA were removed.

Each remaining sample was normalized and variance-stabilized with SCTransform and subjected to PCA, UMAP, and graph-based clustering to obtain preliminary clusters used as input to contamination estimation. Seurat objects were converted to SingleCellExperiment objects and processed with DecontX (celda package)^61^ using the per-sample Seurat clusters as initial labels, and cells with estimated contamination ≥0.5 were excluded. The decontaminated, QC-filtered samples were then integrated using an SCT-based RPCA integration workflow. A shared low-dimensional representation was obtained by running PCA on the integrated assay followed by UMAP, nearest-neighbor graph construction, and clustering at resolution 0.5 (**Figure S12A**).

#### Major cell-type annotation and tumor-cell subset

Integrated clusters were annotated to broad cell types by inspecting canonical marker genes for fibroblasts (*COL3A1*, *LUM*, *DCN*), T cells (*PTPRC*, *CD3D*, *IL7R*, *GZMA*), B cells (*PTPRC*, *CD79A*, *MS4A1*, *SDC1*), NK cells (*PTPRC*, *NKG7*, *GNLY*, *KLRD1*), myeloid cells (*PTPRC*, *CD14*, *LYZ*, *S100A8*), endothelial cells (*PECAM1*, *VWF*, *CD34*, *TIE1*), pancreatic endocrine cells (*INS*, *GCG*, *SST*, *GHRL*), and pancreatic exocrine cells (*CPA1*, *AMY2A*, *KRT19*, *SOX9*) on the integrated UMAP embedding. Clusters exhibiting clear marker signatures were labeled as Endocrine, Acinar, Duct, Monocytes, Endothelial, or Fibroblasts (**Figure S12B**); all remaining clusters were considered tumor epithelial cells and selected as the tumor cell subset used for subsequent analyses. Tumor-specific embeddings were generated and cells were re-clustered at resolution 0.5.

#### Three-family tumor subtyping using NMF signatures

To assign tumors to Classical, Hybrid, and Basal transcriptional families, we used the published NMF signatures from Chan-Seng-Yue et al. (2020).^45^ The four NMF components most relevant to the Classical–Basal continuum were imported and converted into gene sets (**Table S10**), intersected with the expressed gene universe, and summarized per cell as the mean log-normalized expression of the genes in each signature. Per-cell “classical” and “basal” signals were defined as Sig1+Sig6 and Sig2+Sig10, respectively, each normalized by the total signal across all four signatures to obtain fractions “p_classical” and “p_basal”. Cells were then assigned to three tumor families using the following thresholds: Classical if p_classical ≥ 0.60 and p_basal ≤ 0.30, Basal if p_basal ≥ 0.60 and p_classical ≤ 0.30, and Hybrid if both p_classical and p_basal were at least 0.30.

#### TF activity inference and PDX1 activity

For tumor cells, transcription factor activities were inferred from log-normalized RNA using VIPER.^53,54^ We constructed two regulon collections: (i) the human DoRothEA^62^ regulon filtered to confidence levels A–C, converted to a regulon object, and (ii) the CollecTRI^63^ TF–target network. VIPER was run separately on both regulons (minimum regulon size 5), and TF activity matrices from DoRothEA and CollecTRI were merged into a union activity matrix, prioritizing non-redundant TFs when necessary. Activity scores were z-scaled before use in downstream models.

#### Hurdle models relating gene expression to PDX1 activity

To quantify gene-level associations between PDX1 activity and tumor-cell expression, we used a two-part hurdle framework analogous to that employed for the snRNA-seq datasets. We defined for each gene a binary detection indicator (det = 1 if expression > 0, else 0) and a continuous level (SCTransform value) among cells with detection. For each gene with at least 25 expressing cells, we fit a logistic regression for detection and a linear regression for expression among positives, using standardized PDX1 activity as the main predictor and adjusting for technical and biological covariates: the number of detected genes per cell and fixed effects for tumor cluster and sample.

From each model we extracted the PDX1 coefficient and *P*-value, reporting for the detection component the log-odds ratio (logOR_det), odds ratio (OR_det), and *P*-value (p_det), and for the continuous component the slope (beta_cont) and *P*-value (p_cont). Genes with modeling errors or insufficient positive cells were removed. Per-gene *P*-values were combined using Fisher’s method to obtain an overall *P*-value (p_combined), and Benjamini–Hochberg FDRs were computed for detection, continuous, and combined components (FDR_det, FDR_cont, FDR_comb). The full hurdle-model results are reported in **Table S11**.

#### GSEA rank files and pathway enrichment analysis

For gene set enrichment analysis, we converted the detection and continuous *P*-values to signed z-scores using the sign of the corresponding coefficients and combined them into 2:1 (detection:continuous) weighted combined scores. Extreme z-scores were clipped to reasonable bounds to stabilize the tails, and the pre-ranked gene list containing gene symbols and their associated z-scores were used for Gene Set Enrichment Analysis (GSEA).^64^ GSEA was run externally with WebGestalt (https://www.webgestalt.org/#) against GO: Biological Process^39,40^, MSigDB Hallmark^41^, and KEGG^42^ pathway collections (**Figures 5C-D**, and **Tables S12-S14**).

## Data Availability

All data used by or produced in the present study are available online either open-access or upon request to the corresponding data access committee (DAC) as detailed in the manuscript.

https://github.com/hoskinsjw/PDX1_post-GWAS_study

https://github.com/hoskinsjw/Omics_data_and_annotations_for_Zhong_et_al_2025

https://explore.data.humancellatlas.org/projects/b3938158-4e8d-4fdb-9e13-9e94270dde16

https://ega-archive.org/datasets/EGAD00010001811

## Declaration of interests

The authors declare no competing interests.

## Acknowledgements

This research was supported by the Intramural Research Program of the National Institutes of Health (NIH). The contributions of the NIH author(s) are considered Works of the United States Government. The content of this publication does not necessarily reflect the views or policies of the Department of Health and Human Services, nor does mention of trade names, commercial products, or organizations imply endorsement by the U.S. Government.

This study utilized the high-performance computational capabilities of the Biowulf Linux cluster at the NIH, Bethesda, MD, USA (http://biowulf.nih.gov). The authors acknowledge the research contributions of the Cancer Genomics Research Laboratory for their expertise, execution, and support of this research in the areas of project planning, wet laboratory processing of specimens, and bioinformatics analysis of generated data. We also thank participants and clinical coordinators participating in the Pancreatic Cancer Cohort Consortium, Pancreatic Cancer Case-Control Consortium and the UK Biobank for providing samples for the GWAS studies. This project has been funded in whole or in part with Federal funds from the National Cancer Institute, National Institutes of Health, under NCI Contract No. 75N910D00024. This study was conducted with the support of data which comes from the Ontario Institute for Cancer Research through funding provided by the Government of Ontario.

## Author contributions

J.W.H. (Conceptualization, Formal analysis, Investigation, Methodology, Project administration, Software, Supervision, Visualization, Writing – original draft, Writing – review & editing), T.A.C (Investigation), D.E. (Investigation), E.C. (Investigation), M.M. (Investigation), A.O. (Writing – review & editing), I.C. (Resources, Writing – review & editing), J.Z. (Writing – review & editing), M.B.P. (Writing – review & editing), G.P. (Writing – review & editing), H.E.A. (Writing – review & editing), K.E.C. (Writing – review & editing), L.T.A. (Conceptualization, Funding acquisition, Supervision, Writing – review & editing).

## Data and code availability

No new programs or packages were created for this study, but scripts and relevant input files used to run analyses are available in the following GitHub repository (https://github.com/hoskinsjw/PDX1_post-GWAS_study). Browser track files for the pancreatic epigenomic annotations from Zhong et al. (2026)^30^ are available in the following GitHub repository (https://github.com/hoskinsjw/Omics_data_and_annotations_for_Zhong_et_al_2025). The snRNA-seq pancreatic datasets from Tosti et al. (2021)^36^ may be downloaded through the Human Cell Atlas Data Explorer (https://explore.data.humancellatlas.org/projects/b3938158-4e8d-4fdb-9e13-9e94270dde16). The PDAC scRNA-seq dataset from Chan-Seng-Yue et al. (2020)^45^ is available upon request to the Ontario Institute for Cancer Research Data Access Committee (OICR-DAC) through the European Genome-Phenome Archive (https://ega-archive.org/datasets/EGAD00010001811).

## Consortia Authors from the Pancreatic Cancer Cohort Consortium and Pancreatic Cancer Case-Control Consortium (PanScan/PanC4)

Jun Zhong^1^, Nina Afshar^2,3^, Demetrius Albanes^4^, Gabriella Andreotti^5^, Samuel O Antwi^6^, Alan A Arslan^7^, William R Bamlet^8^, Laura Beane-Freeman^5^, Sonja I Berndt^5^, Lea Bouteille^9^, Paige M Bracci^10^, Daniele Campa^11^, Federico Canzian^12^, Stephen J Chanock^13^, Yu Chen^14^, Charles C Chung^15^, Mengmeng Du^16^, A. Heather Eliassen^17,18,19^, Steven Gallinger^20^, J. Michael Gaziano^21,22^, Jeanine Genkinger^23^, Michael Goggins^24^, Phyllis J Goodman^25^, Christopher A Haiman^26^, Manal Hassan^27^, Belynda Hicks^15^, Rayjean J Hung^28^, Amy Hutchinson^15^, Verena Katzke^29^, Manolis Kogevinas^30,31^, Charles Kooperberg^32^, Peter Kraft^33^, Robert C Kurtz^34^, I-Min Lee^35,17^, Loic LeMarchand^36^, Núria Malats^37^, Roger Milne^2,3,38^, Steven C Moore^4^, Rachel E Neale^39^, Kimmie Ng^40^, Ann L Oberg^8^, Irene Orlow^41^, Alpa V Patel^42^, Ulrike Peters^43^, Miquel Porta^44^, Kari G Rabe^45^, Francisco X Real^46,47^, Harvey A. Risch^48^, Nathaniel Rothman^13^, Paul Scheet^49^, Howard D Sesso^21,17^, Veronica W Setiawan^50^, Xiao-Ou Shu^51^, Debra Silverman^13^, Mingyang Song^17,52,53^, Meir J Stampfer^17,18,19^, Melissa C Southey^2,3,38^, Connie Thurman^17^, Geoffrey S Tobias^13^, Caroline Um^42^, Kala Visvanathan^54,55^, Nicolas Wentzensen^13^, Jean Wactawski-Wende^56^, Herbert Yu^57^, Chen Yuan^40^, Wei Zheng^51^, Brian M Wolpin^40^, Rachael Z Stolzenberg-Solomon^13^, Alison P. Klein^58,59^, Laufey T Amundadottir^1^

^1^Laboratory of Translational Genomics, Division of Cancer Epidemiology and Genetics, National Cancer Institute, National Institutes of Health, Bethesda, MD, USA, ^2^Cancer Epidemiology Division, Cancer Council Victoria, Melbourne, VIC, Australia, ^3^Centre for Epidemiology and Biostatistics, Melbourne School of Population and Global Health, The University of Melbourne, Melbourne, VIC, Australia, ^4^Metabolic Epidemiology Branch, Division of Cancer Epidemiology and Genetics, National Cancer Institute, National Institutes of Health, Bethesda, MD, USA, ^5^Occupational and Environmental Epidemiology Branch, Division of Cancer Epidemiology and Genetics, National Cancer Institute, National Institutes of Health, Bethesda, MD, USA, ^6^Department of Quantitative Health Sciences, Mayo Clinic College of Medicine, Jacksonville, FL, USA, ^7^Departments of Obstetrics and Gynecology and Population Health, NYU Grossman School of Medicine, NYU Perlmutter Comprehensive Cancer Center, New York, NY, USA, ^8^Department of Quantitative Health Sciences, Mayo Clinic College of Medicine, Rochester, MN, USA, ^9^Paris-Saclay University, UVSQ, Inserm, Gustave Roussy, CESP, Villejuif, France, ^10^Department of Epidemiology and Biostatistics, University of California San Francisco, San Francisco, CA, USA, ^11^Unit of Genetics., Department of Biology, University of Pisa, Pisa, Italy, ^12^Genomic Epidemiology Group, German Cancer Research Center (DKFZ), Heidelberg, Germany, ^13^Division of Cancer Epidemiology and Genetics, National Cancer Institute, National Institutes of Health, Bethesda, MD, USA, ^14^Department of Population Health, NYU Grossman School of Medicine, NYU Perlmutter Comprehensive Cancer Center, New York, NY, USA, ^15^Cancer Genomics Research Laboratory, Frederick National Lab for Cancer Research, Frederick, MD, USA, ^16^Department of Epidemiology and Biostatistics, Memorial Sloan Kettering Cancer Center,, New York, NY, USA, ^17^Department of Epidemiology, Harvard T.H. Chan School of Public Health, Boston, MA, USA, ^18^Channing Division of Network Medicine, Department of Medicine, Brigham and Women’s Hospital and Harvard Medical School, Boston, MA, USA, ^19^Department of Nutrition, Harvard T. H. Chan School of Public Health, Boston, MA, USA, ^20^Lunenfeld-Tanenbaum Research Institute, Sinai Health System and University of Toronto,, Toronto, Ontario, Canada, ^21^Division of Preventive Medicine, Brigham and Women’s Hospital, Boston, MA, USA, ^22^Division of Aging, Brigham and Women’s Hospital, Boston, MA, USA, ^23^Department of Epidemiology, Columbia University, New York, NY, USA, ^24^Department of Pathology, Johns Hopkins School of Medicine, Baltimore, MD, USA, ^25^SWOG Statistical Center, Fred Hutchinson Cancer Center, Seattle, WA, USA, ^26^Department of Preventive Medicine, Keck School of Medicine, University of Southern California, Los Angeles, CA, USA, ^27^Department of Epidemiology, Cancer Prevention, The University of Texas MD Anderson Cancer Center, Houston, TX, USA, ^28^Prosserman Centre for Population Health Research, Lunenfeld-Tanenbaum Research Institute, Sinai Health System, Toronto, Ontario, Canada, ^29^Division of Cancer Epidemiology, German Cancer Research Center (DKFZ), Heidelberg, Germany, ^30^Environment and Health over the Lifecourse Program, ISGlobal, Barcelona, Spain, ^31^IMIM (Hospital del Mar Medical Research Institute), Barcelona, Spain, ^32^Division of Public Health Sciences, Fred Hutchinson Cancer Center, Seattle, WA, USA, ^33^Trans-Divisional Research Program (TDRP), Division of Cancer Epidemiology and Genetics, National Cancer Institute, National Institutes of Health, Bethesda, MD, USA, ^34^Gastroenterology, Hepatology, and Nutrition Service, Memorial Sloan Kettering Cancer Center, New York, NY, USA, ^35^Division of Preventive Medicine, Department of Medicine, Brigham and Women’s Hospital, Boston, MA, USA, ^36^Population Sciences in the Pacific Program, Cancer Epidemiology, University of Hawaii Cancer Center, Honolulu, HI, USA, ^37^Genetic and Molecular Epidemiology Group, Spanish National Cancer Research Center (CNIO) and CIBERONC, Madrid, Spain, ^38^Precision Medicine, School of Clinical Sciences at Monash Health, Monash University, Clayton, VIC, Australia, ^39^Department of Population Health, QIMR Berghofer Medical Research Institute, Queensland, Australia, ^40^Department of Medical Oncology, Dana-Farber Cancer Institute, Harvard Medical School, Harvard University, Boston, MA, USA, ^41^Department of Epidemiology and Biostatistics, Memorial Sloan Kettering Cancer Center, New York, NY, USA, ^42^Department of Population Science, American Cancer Society, Atlanta, GA, USA, ^43^Division of Public Health Sciences, Fred Hutchinson Cancer Center, Seattle, WA, USA, ^44^Hospital del Mar Institute of Medical Research (IMIM), Centro de Investigacion Biomedica en Red de Epidemiologia y Salud Publica (CIBERESP), Barcelona, Spain, ^45^Department of Quantitative Health Sciences Research, Mayo Clinic College of Medicine, Rochester, MN, USA, ^46^Epithelial Carcinogenesis Group, Tumor Biology Programme, Spanish National Cancer Research Center (CNIO), Madrid, Spain, ^47^Department of Medicine and Life Sciences, Universitat Pompeu Fabra, Barcelona, Spain, ^48^Department of Chronic Disease Epidemiology, Yale School of Public Health, New Haven, CT, USA, ^49^Dept of Epidemiology, The University of Texas MD Anderson Cancer Center, Houston, TX, USA, ^50^Department of Population and Public Health Sciences, Keck School of Medicine, University of Southern California, Los Angeles, CA, USA, ^51^Division of Epidemiology, Department of Medicine, Vanderbilt Epidemiology Center, Vanderbilt-Ingram Cancer Center, Vanderbilt University School of Medicine, Nashville, TN, USA, ^52^Department of Nutrition, Harvard T.H. Chan School of Public Health, Boston, MA, USA, ^53^Clinical and Translational Epidemiology Unit and Division of Gastroenterology, Massachusetts General Hospital and Harvard Medical School, Boston, MA, USA, ^54^Department of Epidemiology, Johns Hopkins School of Public Health, Baltimore, MD, USA, ^55^Department of Oncology, Sidney Kimmel Comprehensive Cancer Center, Johns Hopkins School of Medicine, Baltimore, MD, USA, ^56^Department of Epidemiology and Environmental Health, University of Buffalo, Buffalo, NY, USA, ^57^Epidemiology Program, University of Hawaii Cancer Center, Honolulu, HI, USA, ^58^Department of Oncology, Sidney Kimmel Comprehensive Cancer Center, Johns Hopkins School of Medicine, Baltimore, MD, USA, ^59^Department of Medicine, Division of Gastroenterology & Hepatology, Johns Hopkins School of Medicine, Baltimore, MD, USA.

## Notes

### Competing Interest Statement

The authors have declared no competing interest.

### Author Declarations

All participating studies have institutional IRB approvals and the NCI part of the study has NIH Special Study IRB approval. The GWAS and eQTL data used in this study were summary data, while the sc/snRNA-seq data were individual-level data that had been de-identified prior to use in this study.

## References

1. Sherman, R.L., Firth, A.U., Henley, S.J., Siegel, R.L., Negoita, S., Sung, H., Kohler, B.A., Anderson, R.N., Cucinelli, J., Scott, S., et al. (2025). Annual Report to the Nation on the Status of Cancer, featuring state-level statistics after the onset of the COVID-19 pandemic. Cancer 131, e35833. 10.1002/cncr.35833.

2. Amundadottir, L., Kraft, P., Stolzenberg-Solomon, R.Z., Fuchs, C.S., Petersen, G.M., Arslan, A.A., Bueno-de-Mesquita, H.B., Gross, M., Helzlsouer, K., Jacobs, E.J., et al. (2009). Genome-wide association study identifies variants in the ABO locus associated with susceptibility to pancreatic cancer. Nat Genet 41, 986–990. 10.1038/ng.429.

3. Petersen, G.M., Amundadottir, L., Fuchs, C.S., Kraft, P., Stolzenberg-Solomon, R.Z., Jacobs, K.B., Arslan, A.A., Bueno-de-Mesquita, H.B., Gallinger, S., Gross, M., et al. (2010). A genome-wide association study identifies pancreatic cancer susceptibility loci on chromosomes 13q22.1, 1q32.1 and 5p15.33. Nat Genet 42, 224–228. 10.1038/ng.522.

4. Wolpin, B.M., Rizzato, C., Kraft, P., Kooperberg, C., Petersen, G.M., Wang, Z., Arslan, A.A., Beane-Freeman, L., Bracci, P.M., Buring, J., et al. (2014). Genome-wide association study identifies multiple susceptibility loci for pancreatic cancer. Nat Genet 46, 994–1000. 10.1038/ng.3052.

5. Childs, E.J., Mocci, E., Campa, D., Bracci, P.M., Gallinger, S., Goggins, M., Li, D., Neale, R.E., Olson, S.H., Scelo, G., et al. (2015). Common variation at 2p13.3, 3q29, 7p13 and 17q25.1 associated with susceptibility to pancreatic cancer. Nat Genet 47, 911–916. 10.1038/ng.3341.

6. Zhang, M., Wang, Z., Obazee, O., Jia, J., Childs, E.J., Hoskins, J., Figlioli, G., Mocci, E., Collins, I., Chung, C.C., et al. (2016). Three new pancreatic cancer susceptibility signals identified on chromosomes 1q32.1, 5p15.33 and 8q24.21. Oncotarget 7, 66328–66343. 10.18632/oncotarget.11041.

7. Klein, A.P., Wolpin, B.M., Risch, H.A., Stolzenberg-Solomon, R.Z., Mocci, E., Zhang, M., Canzian, F., Childs, E.J., Hoskins, J.W., Jermusyk, A., et al. (2018). Genome-wide meta-analysis identifies five new susceptibility loci for pancreatic cancer. Nat Commun 9, 556. 10.1038/s41467-018-02942-5.

8. Tam, V., Patel, N., Turcotte, M., Bosse, Y., Pare, G., and Meyre, D. (2019). Benefits and limitations of genome-wide association studies. Nat Rev Genet 20, 467–484. 10.1038/s41576-019-0127-1.

9. Gallagher, M.D., and Chen-Plotkin, A.S. (2018). The Post-GWAS Era: From Association to Function. Am J Hum Genet 102, 717–730. 10.1016/j.ajhg.2018.04.002.

10. Cano-Gamez, E., and Trynka, G. (2020). From GWAS to Function: Using Functional Genomics to Identify the Mechanisms Underlying Complex Diseases. Front Genet 11, 424. 10.3389/fgene.2020.00424.

11. Jia, J., Bosley, A.D., Thompson, A., Hoskins, J.W., Cheuk, A., Collins, I., Parikh, H., Xiao, Z., Ylaya, K., Dzyadyk, M., et al. (2014). CLPTM1L promotes growth and enhances aneuploidy in pancreatic cancer cells. Cancer Res 74, 2785–2795. 10.1158/0008-5472.CAN-13-3176.

12. Hoskins, J.W., Jia, J., Flandez, M., Parikh, H., Xiao, W., Collins, I., Emmanuel, M.A., Ibrahim, A., Powell, J., Zhang, L., et al. (2014). Transcriptome analysis of pancreatic cancer reveals a tumor suppressor function for HNF1A. Carcinogenesis 35, 2670–2678. 10.1093/carcin/bgu193.

13. Fang, J., Jia, J., Makowski, M., Xu, M., Wang, Z., Zhang, T., Hoskins, J.W., Choi, J., Han, Y., Zhang, M., et al. (2017). Functional characterization of a multi-cancer risk locus on chr5p15.33 reveals regulation of TERT by ZNF148. Nat Commun 8, 15034. 10.1038/ncomms15034.

14. Jermusyk, A., Zhong, J., Connelly, K.E., Gordon, N., Perera, S., Abdolalizadeh, E., Zhang, T., O’Brien, A., Hoskins, J.W., Collins, I., et al. (2021). A 584 bp deletion in CTRB2 inhibits chymotrypsin B2 activity and secretion and confers risk of pancreatic cancer. Am J Hum Genet 108, 1852–1865. 10.1016/j.ajhg.2021.09.002.

15. Zhong, J., Jermusyk, A., Wu, L., Hoskins, J.W., Collins, I., Mocci, E., Zhang, M., Song, L., Chung, C.C., Zhang, T., et al. (2020). A Transcriptome-Wide Association Study Identifies Novel Candidate Susceptibility Genes for Pancreatic Cancer. J Natl Cancer Inst 112, 1003–1012. 10.1093/jnci/djz246.

16. Bodas, C., Felipe, I., Chanez, B., Lafarga, M., López De Maturana, E., Martínez-de-Villarreal, J., Del Pozo, N., Malumbres, M., Vargiu, P., Cayuela, A., et al. (2024). A common CTRB misfolding variant associated with pancreatic cancer risk causes ER stress and inflammation in mice. Preprint at Cancer Biology, 10.1101/2024.07.23.604778 https://doi.org/10.1101/2024.07.23.604778.

17. Connelly, K.E., Hullin, K., Abdolalizadeh, E., Zhong, J., Eiser, D., O’Brien, A., Collins, I., Das, S., Duncan, G., Pancreatic Cancer Cohort, C., et al. (2025). Allelic effects on KLHL17 expression underlie a pancreatic cancer genome-wide association signal at chr1p36.33. Nat Commun 16, 4055. 10.1038/s41467-025-59109-2.

18. Wang, M., and Kaufman, R.J. (2014). The impact of the endoplasmic reticulum protein-folding environment on cancer development. Nat Rev Cancer 14, 581–597. 10.1038/nrc3800.

19. Molero, X., Vaquero, E.C., Flández, M., González, A.M., Ortiz, M.Á., Cibrián-Uhalte, E., Servitja, J.-M., Merlos, A., Juanpere, N., Massumi, M., et al. (2012). Gene expression dynamics after murine pancreatitis unveils novel roles for Hnf1α in acinar cell homeostasis. Gut 61, 1187–1196. 10.1136/gutjnl-2011-300360.

20. Camolotto, S.A., Belova, V.K., and Snyder, E.L. (2018). The role of lineage specifiers in pancreatic ductal adenocarcinoma. J. Gastrointest. Oncol 9, 1005–1013. 10.21037/jgo.2018.05.04.

21. Cobo, I., Martinelli, P., Flández, M., Bakiri, L., Zhang, M., Carrillo-de-Santa-Pau, E., Jia, J., Sánchez-Arévalo Lobo, V.J., Megías, D., Felipe, I., et al. (2018). Transcriptional regulation by NR5A2 links differentiation and inflammation in the pancreas. Nature 554, 533–537. 10.1038/nature25751.

22. Kalisz, M., Bernardo, E., Beucher, A., Maestro, M.A., Del Pozo, N., Millán, I., Haeberle, L., Schlensog, M., Safi, S.A., Knoefel, W.T., et al. (2020). HNF1A recruits KDM6A to activate differentiated acinar cell programs that suppress pancreatic cancer. EMBO J 39, e102808. 10.15252/embj.2019102808.

23. Yang, S., Tang, W., Azizian, A., Gaedcke, J., Ströbel, P., Wang, L., Cawley, H., Ohara, Y., Valenzuela, P., Zhang, L., et al. (2022). Dysregulation of HNF1B/Clusterin axis enhances disease progression in a highly aggressive subset of pancreatic cancer patients. Carcinogenesis 43, 1198–1210. 10.1093/carcin/bgac092.

24. Rao, S.V., Young, L., Cheeseman, D., Flynn, S., Krebs, N., Couturier, D.-L., Mack, S., Brais, R., Temple, J., Smith, A., et al. (2025). Transcription factor switching drives subtype-specific pancreatic cancer. Nat Genet 57, 3016–3026. 10.1038/s41588-025-02389-7.

25. Abel, E.V., Goto, M., Magnuson, B., Abraham, S., Ramanathan, N., Hotaling, E., Alaniz, A.A., Kumar-Sinha, C., Dziubinski, M.L., Urs, S., et al. (2018). HNF1A is a novel oncogene that regulates human pancreatic cancer stem cell properties. Elife 7. 10.7554/eLife.33947.

26. Bailey, P., Chang, D.K., Nones, K., Johns, A.L., Patch, A.-M., Gingras, M.-C., Miller, D.K., Christ, A.N., Bruxner, T.J.C., Quinn, M.C., et al. (2016). Genomic analyses identify molecular subtypes of pancreatic cancer. Nature 531, 47–52. 10.1038/nature16965.

27. Laise, P., Turunen, M., Garcia, A.C., Tomassoni, L., Maurer, H.C., Elyada, E., Schmierer, B., Worley, J., Kesner, J., Tan, X., et al. (2020). Developmental and MAPK-responsive transcription factors drive distinct malignant subtypes and genetic dependencies in pancreatic cancer. Preprint at Cancer Biology, 10.1101/2020.10.27.357269 https://doi.org/10.1101/2020.10.27.357269.

28. Wang, G., Sarkar, A., Carbonetto, P., and Stephens, M. (2020). A simple new approach to variable selection in regression, with application to genetic fine mapping. J R Stat Soc Series B Stat Methodol 82, 1273–1300. 10.1111/rssb.12388.

29. McCreight, A., Cho, Y., Li, R., Nachun, D., Gan, H.-Y., Carbonetto, P., Stephens, M., Denault, W.R.P., and Wang, G. (2025). SuSiE 2.0: improved methods and implementations for genetic fine-mapping and phenotype prediction. Preprint at bioRxiv, 10.1101/2025.11.25.690514 https://doi.org/10.1101/2025.11.25.690514.

30. Zhong, J., O’Brien, A., Patel, M.B., Eiser, D., Mobaraki, M., Collins, I., Wang, L., Guo, K., TruongVo, T., Jermusyk, A., et al. (2026). Large-scale multiomic analysis identifies non-coding somatic driver mutations and nominates ZFP36L2 as a driver gene for pancreatic ductal adenocarcinoma. Gut 75, 563–575. 10.1136/gutjnl-2025-335152.

31. Arda, H.E., Tsai, J., Rosli, Y.R., Giresi, P., Bottino, R., Greenleaf, W.J., Chang, H.Y., and Kim, S.K. (2018). A Chromatin Basis for Cell Lineage and Disease Risk in the Human Pancreas. Cell Syst 7, 310–322.e4. 10.1016/j.cels.2018.07.007.

32. GTEx Consortium (2020). The GTEx Consortium atlas of genetic regulatory effects across human tissues. Science 369, 1318–1330. 10.1126/science.aaz1776.

33. Giambartolomei, C., Vukcevic, D., Schadt, E.E., Franke, L., Hingorani, A.D., Wallace, C., and Plagnol, V. (2014). Bayesian test for colocalisation between pairs of genetic association studies using summary statistics. PLoS Genet 10, e1004383. 10.1371/journal.pgen.1004383.

34. Akerman, I., Tu, Z., Beucher, A., Rolando, D.M.Y., Sauty-Colace, C., Benazra, M., Nakic, N., Yang, J., Wang, H., Pasquali, L., et al. (2017). Human Pancreatic β Cell lncRNAs Control Cell-Specific Regulatory Networks. Cell Metab 25, 400–411. 10.1016/j.cmet.2016.11.016.

35. Ebrahim, N., Shakirova, K., and Dashinimaev, E. (2022). PDX1 is the cornerstone of pancreatic beta-cell functions and identity. Front Mol Biosci 9, 1091757. 10.3389/fmolb.2022.1091757.

36. Tosti, L., Hang, Y., Debnath, O., Tiesmeyer, S., Trefzer, T., Steiger, K., Ten, F.W., Lukassen, S., Ballke, S., Kuhl, A.A., et al. (2021). Single-Nucleus and In Situ RNA-Sequencing Reveal Cell Topographies in the Human Pancreas. Gastroenterology 160, 1330–1344 e11. 10.1053/j.gastro.2020.11.010.

37. Zhang, F., Wu, Y., and Tian, W. (2019). A novel approach to remove the batch effect of single-cell data. Cell Discov 5, 46. 10.1038/s41421-019-0114-x.

38. Uhlén, M., Fagerberg, L., Hallström, B.M., Lindskog, C., Oksvold, P., Mardinoglu, A., Sivertsson, Å., Kampf, C., Sjöstedt, E., Asplund, A., et al. (2015). Proteomics. Tissue-based map of the human proteome. Science 347, 1260419. 10.1126/science.1260419.

39. Ashburner, M., Ball, C.A., Blake, J.A., Botstein, D., Butler, H., Cherry, J.M., Davis, A.P., Dolinski, K., Dwight, S.S., Eppig, J.T., et al. (2000). Gene ontology: tool for the unification of biology. The Gene Ontology Consortium. Nat Genet 25, 25–29. 10.1038/75556.

40. Gene Ontology Consortium (2026). The Gene Ontology knowledgebase in 2026. Nucleic Acids Res 54, D1779–D1792. 10.1093/nar/gkaf1292.

41. Liberzon, A., Birger, C., Thorvaldsdóttir, H., Ghandi, M., Mesirov, J.P., and Tamayo, P. (2015). The Molecular Signatures Database (MSigDB) hallmark gene set collection. Cell Syst 1, 417–425. 10.1016/j.cels.2015.12.004.

42. Kanehisa, M., and Goto, S. (2000). KEGG: kyoto encyclopedia of genes and genomes. Nucleic Acids Res 28, 27–30. 10.1093/nar/28.1.27.

43. Jonckheere, N., Skrypek, N., and Van Seuningen, I. (2010). Mucins and pancreatic cancer. Cancers (Basel) 2, 1794–1812. 10.3390/cancers2041794.

44. Nishimon, R., Yoshida, K., Sanuki, F., Nakashima, Y., Miyake, T., Sato, T., Tomiyama, Y., Nishina, S., Moriya, T., Shiotani, A., et al. (2023). Pancreatic ductal adenocarcinoma with acinar-to-ductal metaplasia-like cancer cells shows increased cellular proliferation. Pancreatology 23, 811–817. 10.1016/j.pan.2023.08.007.

45. Chan-Seng-Yue, M., Kim, J.C., Wilson, G.W., Ng, K., Figueroa, E.F., O’Kane, G.M., Connor, A.A., Denroche, R.E., Grant, R.C., McLeod, J., et al. (2020). Transcription phenotypes of pancreatic cancer are driven by genomic events during tumor evolution. Nat Genet 52, 231–240. 10.1038/s41588-019-0566-9.

46. Guerra, C., Schuhmacher, A.J., Cañamero, M., Grippo, P.J., Verdaguer, L., Pérez-Gallego, L., Dubus, P., Sandgren, E.P., and Barbacid, M. (2007). Chronic pancreatitis is essential for induction of pancreatic ductal adenocarcinoma by K-Ras oncogenes in adult mice. Cancer Cell 11, 291–302. 10.1016/j.ccr.2007.01.012.

47. Song, S.Y., Gannon, M., Washington, M.K., Scoggins, C.R., Meszoely, I.M., Goldenring, J.R., Marino, C.R., Sandgren, E.P., Coffey, R.J., Wright, C.V., et al. (1999). Expansion of Pdx1-expressing pancreatic epithelium and islet neogenesis in transgenic mice overexpressing transforming growth factor alpha. Gastroenterology 117, 1416–1426. 10.1016/s0016-5085(99)70292-1.

48. Rooman, I., De Medts, N., Baeyens, L., Lardon, J., De Breuck, S., Heimberg, H., and Bouwens, L. (2006). Expression of the Notch signaling pathway and effect on exocrine cell proliferation in adult rat pancreas. Am J Pathol 169, 1206–1214. 10.2353/ajpath.2006.050926.

49. Miyatsuka, T., Kaneto, H., Shiraiwa, T., Matsuoka, T., Yamamoto, K., Kato, K., Nakamura, Y., Akira, S., Takeda, K., Kajimoto, Y., et al. (2006). Persistent expression of PDX-1 in the pancreas causes acinar-to-ductal metaplasia through Stat3 activation. Genes Dev 20, 1435–1440. 10.1101/gad.1412806.

50. Roy, N., Takeuchi, K.K., Ruggeri, J.M., Bailey, P., Chang, D., Li, J., Leonhardt, L., Puri, S., Hoffman, M.T., Gao, S., et al. (2016). PDX1 dynamically regulates pancreatic ductal adenocarcinoma initiation and maintenance. Genes Dev 30, 2669–2683. 10.1101/gad.291021.116.

51. Liu, Y., MacDonald, R.J., and Swift, G.H. (2001). DNA binding and transcriptional activation by a PDX1.PBX1b.MEIS2b trimer and cooperation with a pancreas-specific basic helix-loop-helix complex. J Biol Chem 276, 17985–17993. 10.1074/jbc.M100678200.

52. Rotti, P.G., Yi, Y., Gasser, G., Yuan, F., Sun, X., Apak-Evans, I., Wu, P., Liu, G., Choi, S., Reeves, R., et al. (2024). CFTR represses a PDX1 axis to govern pancreatic ductal cell fate. iScience 27, 111393. 10.1016/j.isci.2024.111393.

53. Alvarez, M.J., Shen, Y., Giorgi, F.M., Lachmann, A., Ding, B.B., Ye, B.H., and Califano, A. (2016). Functional characterization of somatic mutations in cancer using network-based inference of protein activity. Nat Genet 48, 838–847. 10.1038/ng.3593.

54. Ding, H., Douglass, E.F., Sonabend, A.M., Mela, A., Bose, S., Gonzalez, C., Canoll, P.D., Sims, P.A., Alvarez, M.J., and Califano, A. (2018). Quantitative assessment of protein activity in orphan tissues and single cells using the metaVIPER algorithm. Nat Commun 9, 1471. 10.1038/s41467-018-03843-3.

55. Hoskins, J.W., Chung, C.C., O’Brien, A., Zhong, J., Connelly, K., Collins, I., Shi, J., and Amundadottir, L.T. (2021). Inferred expression regulator activities suggest genes mediating cardiometabolic genetic signals. PLoS Comput Biol 17, e1009563. 10.1371/journal.pcbi.1009563.

56. Hoskins, J.W., Christensen, T.A., and Amundadottir, L.T. (2023). Master regulator activity QTLprotocol to implicate regulatory pathways potentially mediating GWAS signals using eQTL data. STAR Protoc 4, 102362. 10.1016/j.xpro.2023.102362.

57. Khetan, S., Carroll, B.S., and Bulyk, M.L. (2025). Multiple overlapping binding sites determine transcription factor occupancy. Nature 646, 1001–1011. 10.1038/s41586-025-09472-3.

58. Pollak, M. (2008). Insulin and insulin-like growth factor signalling in neoplasia. Nat Rev Cancer 8, 915–928. 10.1038/nrc2536.

59. Gallagher, E.J., and LeRoith, D. (2011). Minireview: IGF, Insulin, and Cancer. Endocrinology 152, 2546–2551. 10.1210/en.2011-0231.

60. Chang, C.C., Chow, C.C., Tellier, L.C., Vattikuti, S., Purcell, S.M., and Lee, J.J. (2015). Second-generation PLINK: rising to the challenge of larger and richer datasets. Gigascience 4, 7. 10.1186/s13742-015-0047-8.

61. Wang, Z., Yang, S., Koga, Y., Corbett, S.E., Shea, C.V., Johnson, W.E., Yajima, M., and Campbell, J.D. (2022). Celda: a Bayesian model to perform co-clustering of genes into modules and cells into subpopulations using single-cell RNA-seq data. NAR Genom Bioinform 4, lqac066. 10.1093/nargab/lqac066.

62. Garcia-Alonso, L., Holland, C.H., Ibrahim, M.M., Turei, D., and Saez-Rodriguez, J. (2019). Benchmark and integration of resources for the estimation of human transcription factor activities. Genome Res 29, 1363–1375. 10.1101/gr.240663.118.

63. Müller-Dott, S., Tsirvouli, E., Vazquez, M., Ramirez Flores, R.O., Badia-I-Mompel, P., Fallegger, R., Türei, D., Lægreid, A., and Saez-Rodriguez, J. (2023). Expanding the coverage of regulons from high-confidence prior knowledge for accurate estimation of transcription factor activities. Nucleic Acids Res 51, 10934–10949. 10.1093/nar/gkad841.

64. Subramanian, A., Tamayo, P., Mootha, V.K., Mukherjee, S., Ebert, B.L., Gillette, M.A., Paulovich, A., Pomeroy, S.L., Golub, T.R., Lander, E.S., et al. (2005). Gene set enrichment analysis: a knowledge-based approach for interpreting genome-wide expression profiles. Proc Natl Acad Sci U S A 102, 15545–15550. 10.1073/pnas.0506580102.

65. Street, K., Risso, D., Fletcher, R.B., Das, D., Ngai, J., Yosef, N., Purdom, E., and Dudoit, S. (2018). Slingshot: cell lineage and pseudotime inference for single-cell transcriptomics. BMC Genomics 19, 477. 10.1186/s12864-018-4772-0.

