## Supplemental Figures S1-S12 for "Characterization of a pancreatic cancer GWAS signal suggests PDX1 buffers stress in the exocrine pancreas"

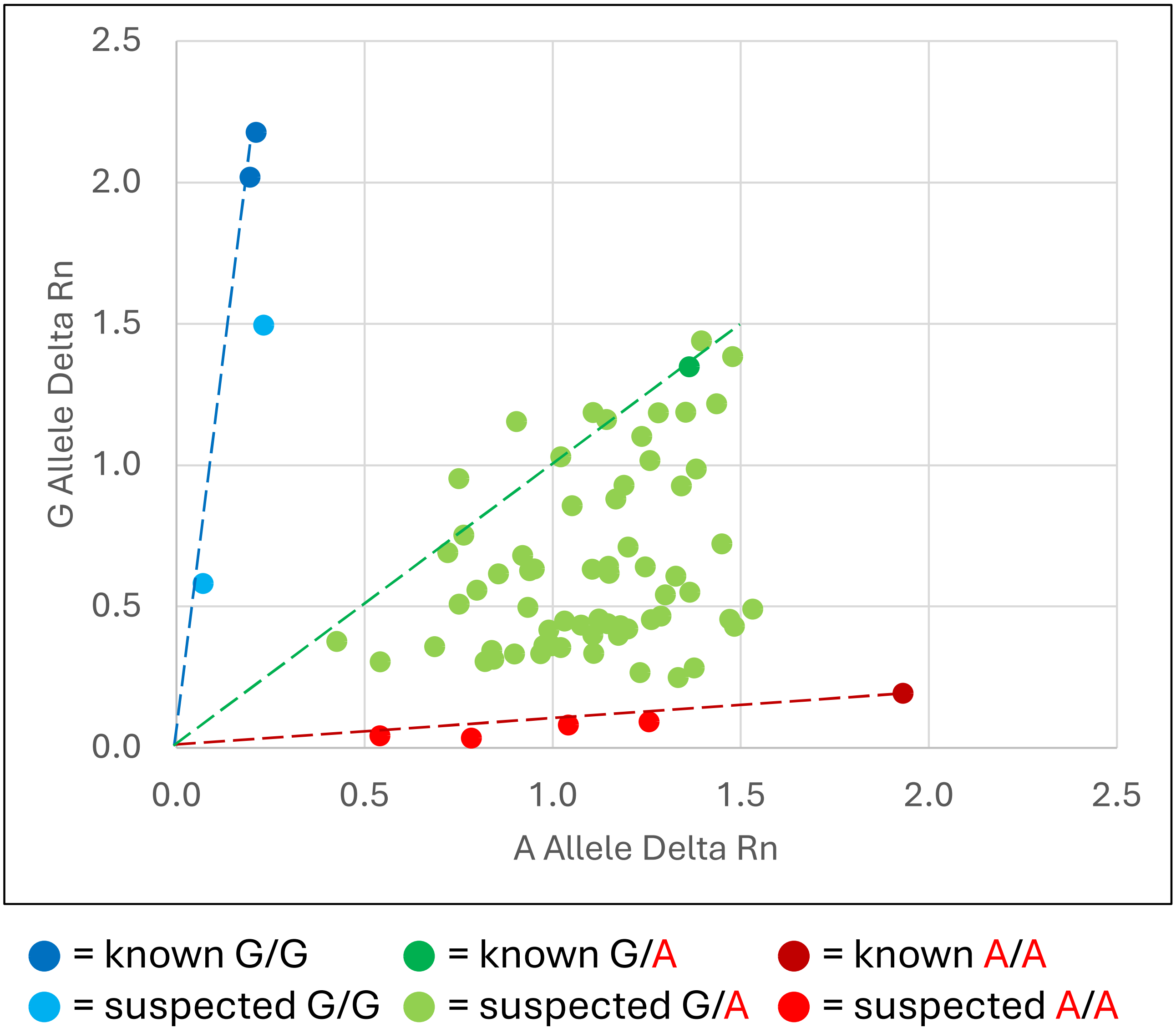


**Figure S1: Genotyping rs9581943 in CRISPR/Cas9-edited Panc 05.04 clones.** Scatter plot summarizing the genotyping qPCR results for the rs9581943 CRISPR/Cas9 edited Panc 05.04 clones using a dual-color genotyping TaqMan assay. Blue dots indicate likely GG clones, red dots indicate likely AA clones, and green dots indicate uncertain or likely unedited GA clones.

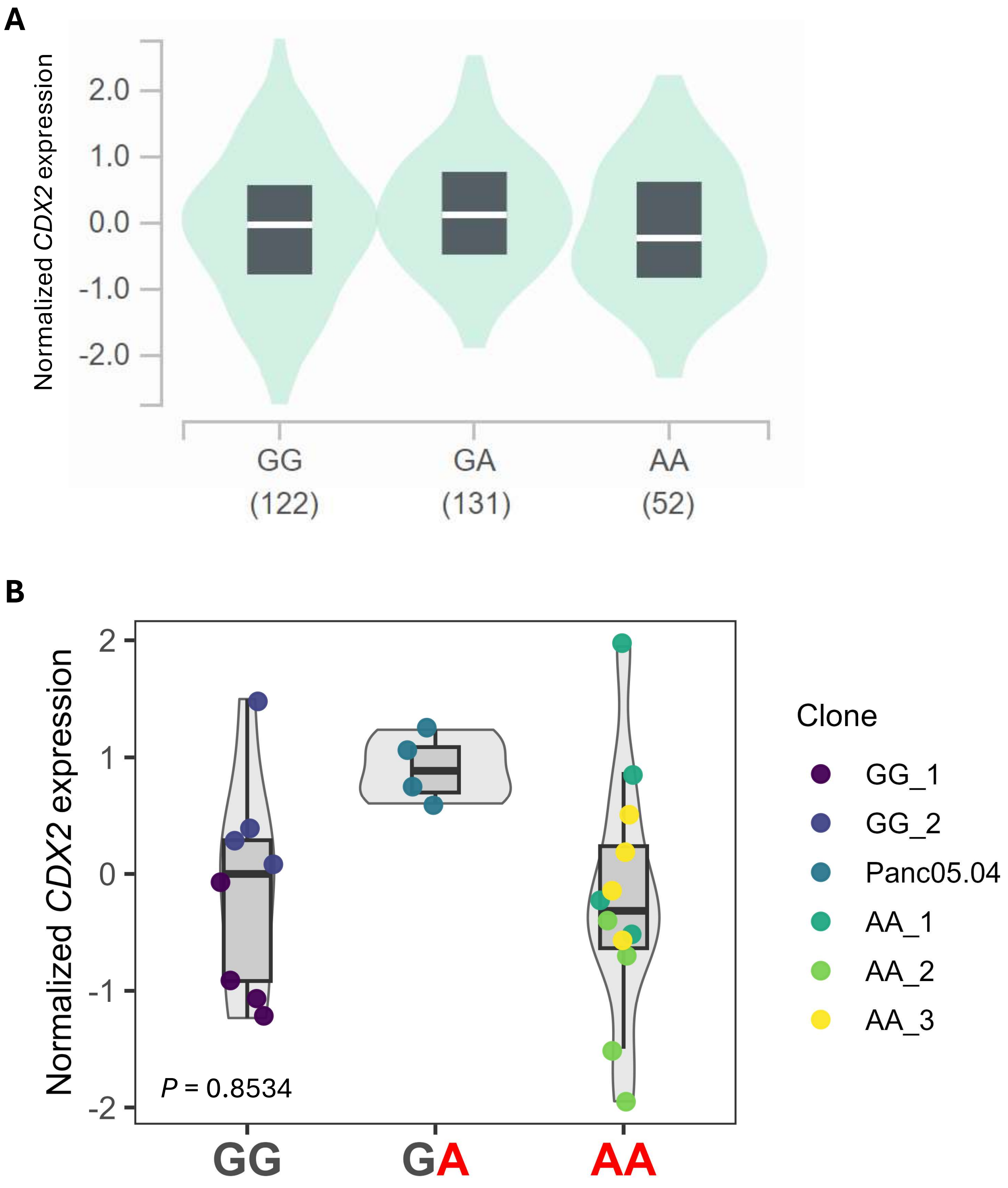


**Figure S2: rs9581943 genotype does not significantly associate with *CDX2* expression. (A)** Violin plot demonstrating the GTEx v8 pancreas eQTL association (or lack thereof) between rs9581943 genotype and *CDX2* normalized expression. **(B)** Violin plot demonstrating the association (or lack thereof) between rs9581943 genotype in CRISPR/Cas9-edited Panc 05.04 clones and *CDX2* normalized expression.

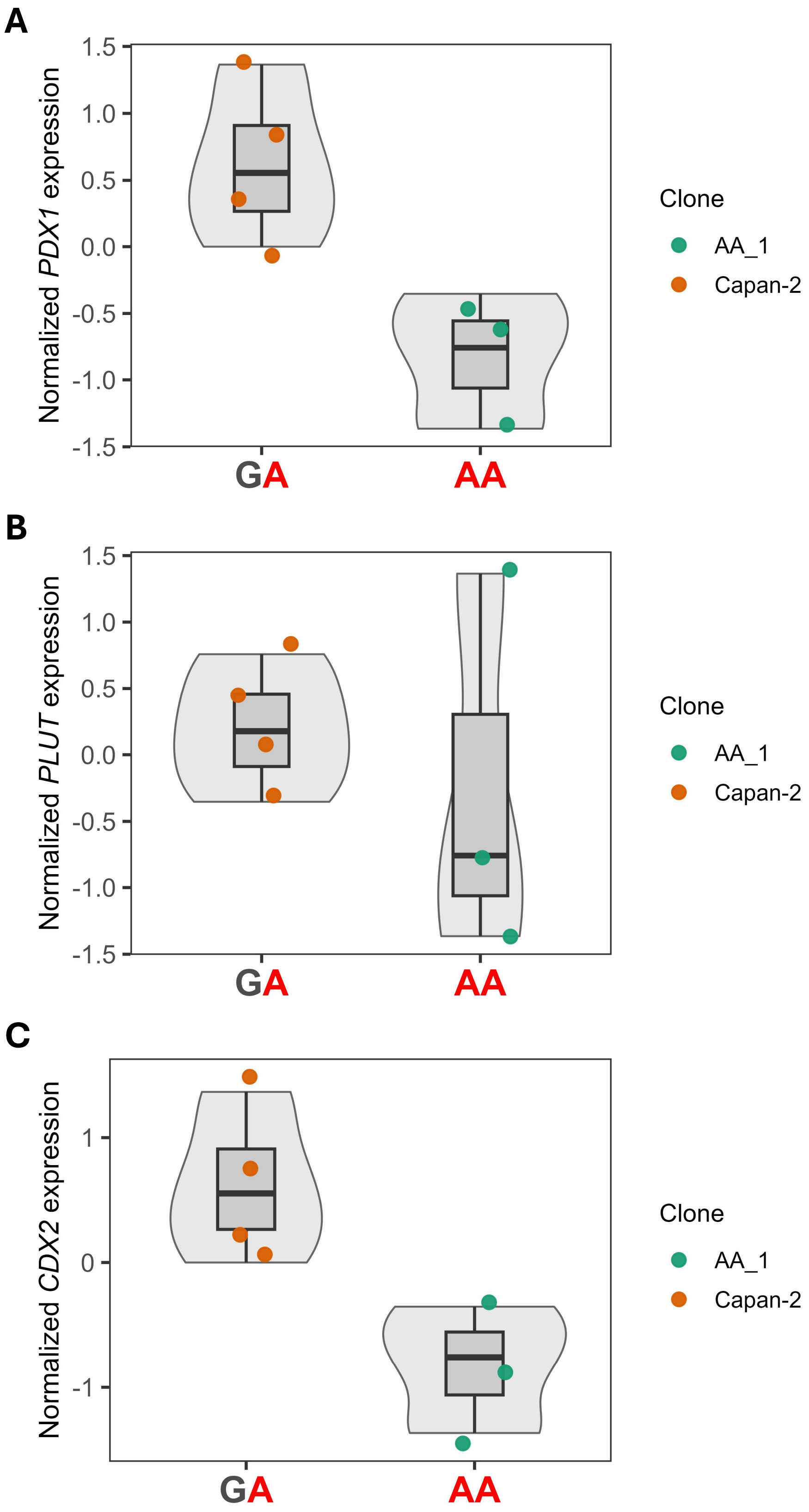


**Figure S3: Expression trends for *PDX1*, *PLUT,* and *CDX2* in a single CRISPR/Cas9 edited Capan-2 clone. (A-C)** Violin plots demonstrating the trends between rs9581943 genotype of a single CRISPR/Cas9-edited Capan-2 clone and *PDX1* **(A)**, *PLUT* **(B)**, or *CDX2* **(C)** normalized expression.

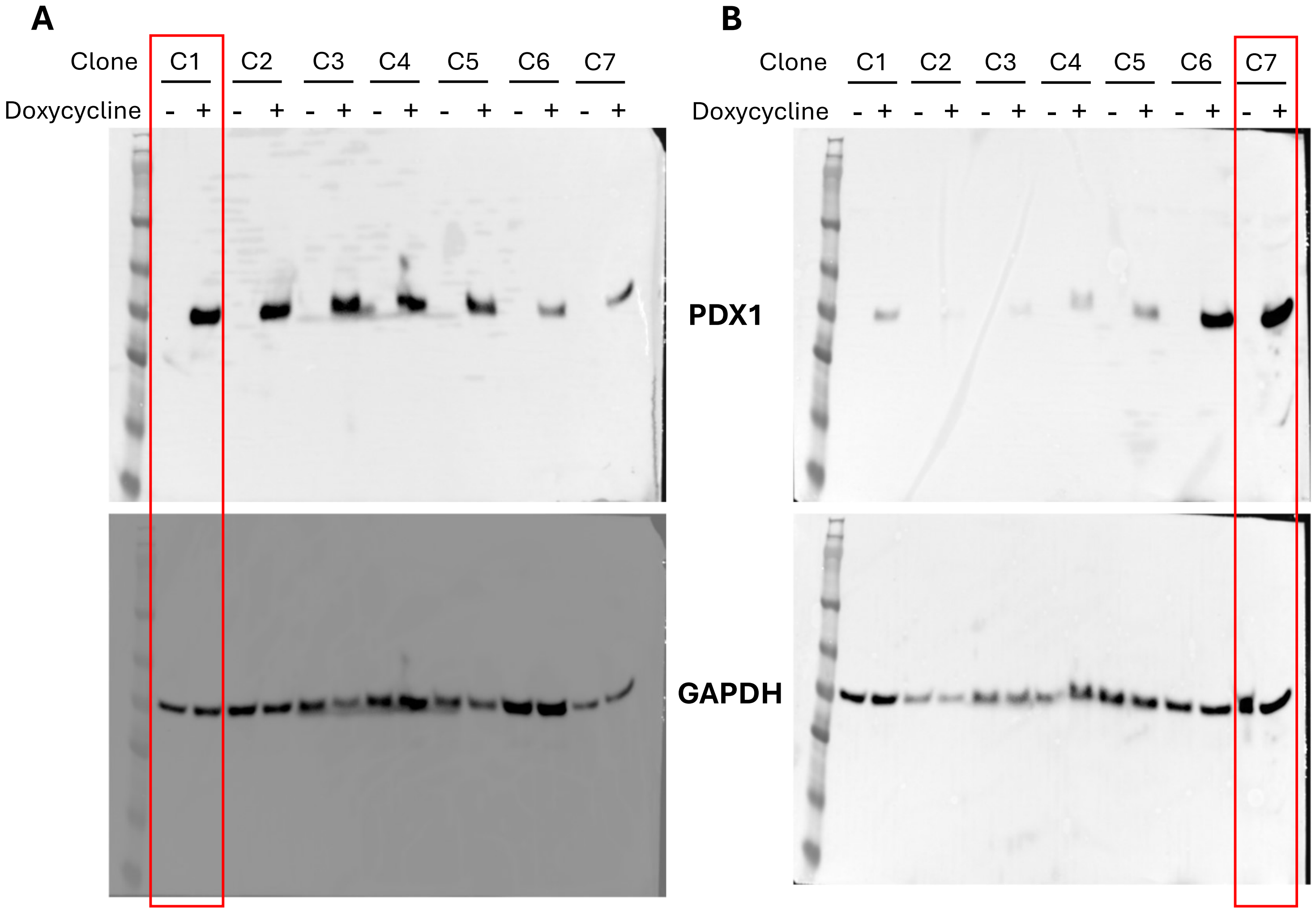


**Figure S4: Western blot indicates doxycycline-induction of PDX1 expression in tested clones.** Western blot analysis of PDX1-inducible clones derived from MIA PaCa-2 **(A)** or PANC-1 **(B)** cell lines +/- 100ng/mL doxycycline induction for 24 hours. GAPDH was used as the loading control. The red rectangles indicate the clones taken forward for downstream analyses of PDX1 overexpression phenotypes.

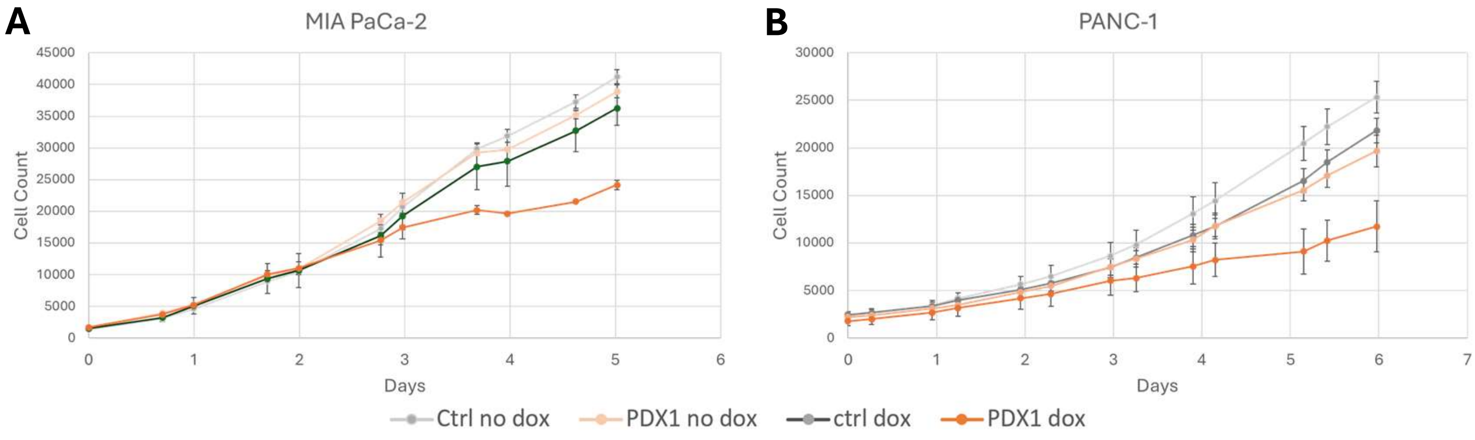


**Figure S5: Induced PDX1 overexpression decreases proliferation in culture.** Cell growth curves for control and PDX1-inducible MIA PaCa-2 **(A)** and PANC-1 **(B)** derived clones cultured +/- 100 ng/mL doxycycline. Doxycycline induction began at the day 0 timepoint. Cell counting was performed with the Lionheart plate reader on cultures in triplicate. Error bars represent the standard error of the mean (SEM).

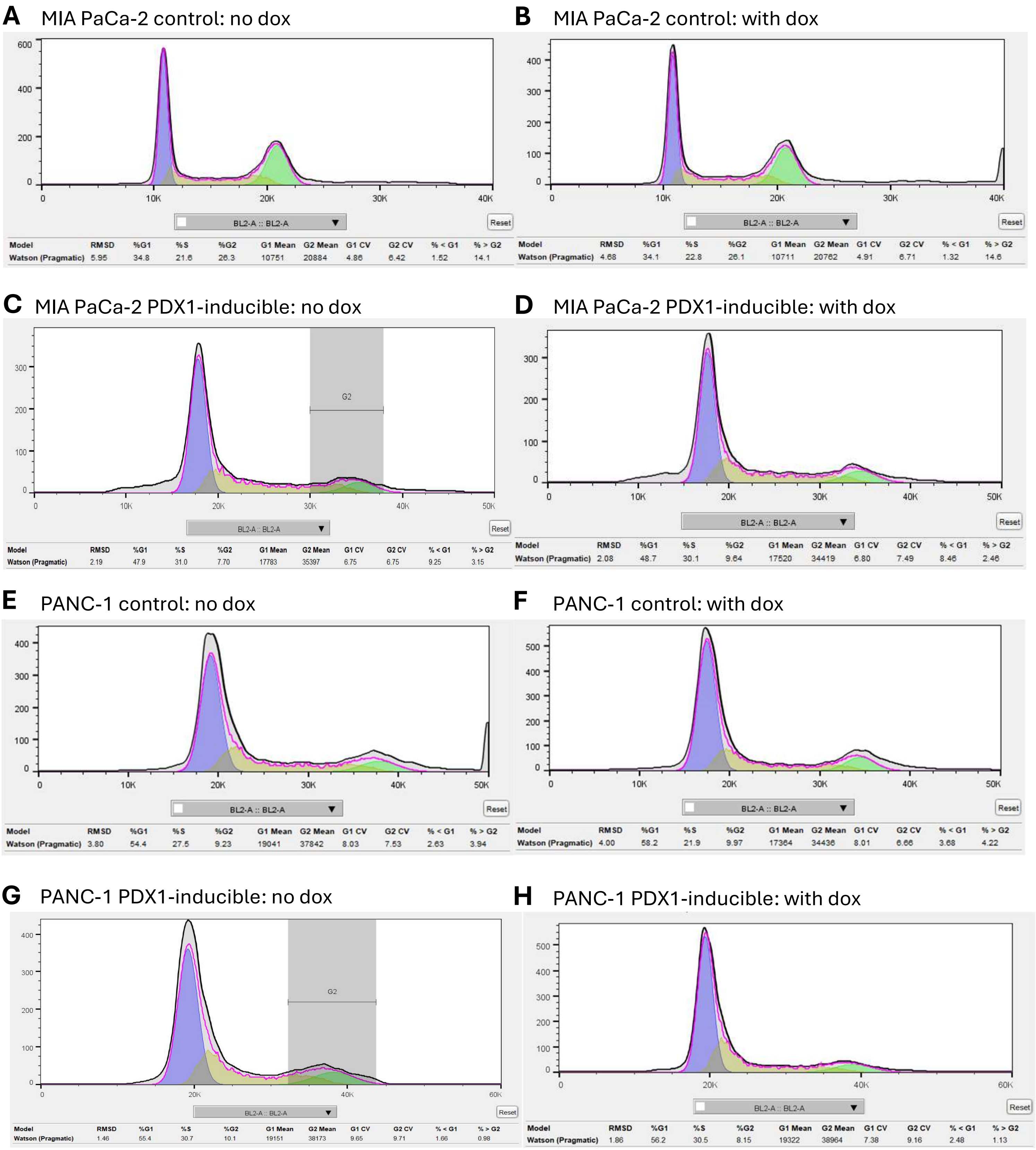


**Figure S6: Induced PDX1 overexpression does not obviously affect the cell cycle in culture.** Representative cell cycle profiles of control **(A-B, E-F)** and PDX-1 inducible **(C-D, G-H)** MIA-PaCa-2 **(A-D)** and PANC-1 **(E-H)** clones grown for four days with **(B, D, F, H)** or without **(A, C, E, G)** 100 ng/mL doxycycline. Cells were fixed and stained with propidium iodide as the DNA stain, and cell cycle profiles (DNA content histograms) were generated by flow cytometry. Cell cycle phase proportions were estimated with ModFit LT software using the Watson (Pragmatic) algorithm and are reported below each curve.

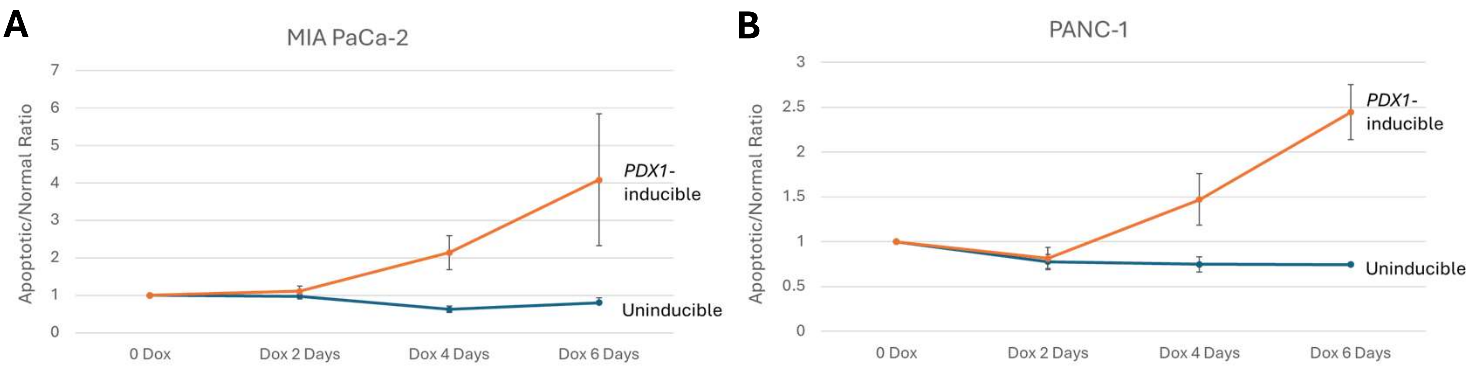


**Figure S7: Induced PDX1 overexpression increases apoptosis in culture.** Apoptosis curves indicating the ratio of apoptotic (Annexin V and propidium iodide positive) to living (Annexin V and propidium iodide negative) cells over time with or without PDX1 induction by 100 ng/mL doxycycline for control and PDX1-inducible MIA PaCa-2 **(A)** and PANC-1 **(B)** derived clones. Cell counts were obtained by flow cytometry and normalized to day 0 timepoints. Error bars represent the standard error of the mean for three independent experiments.

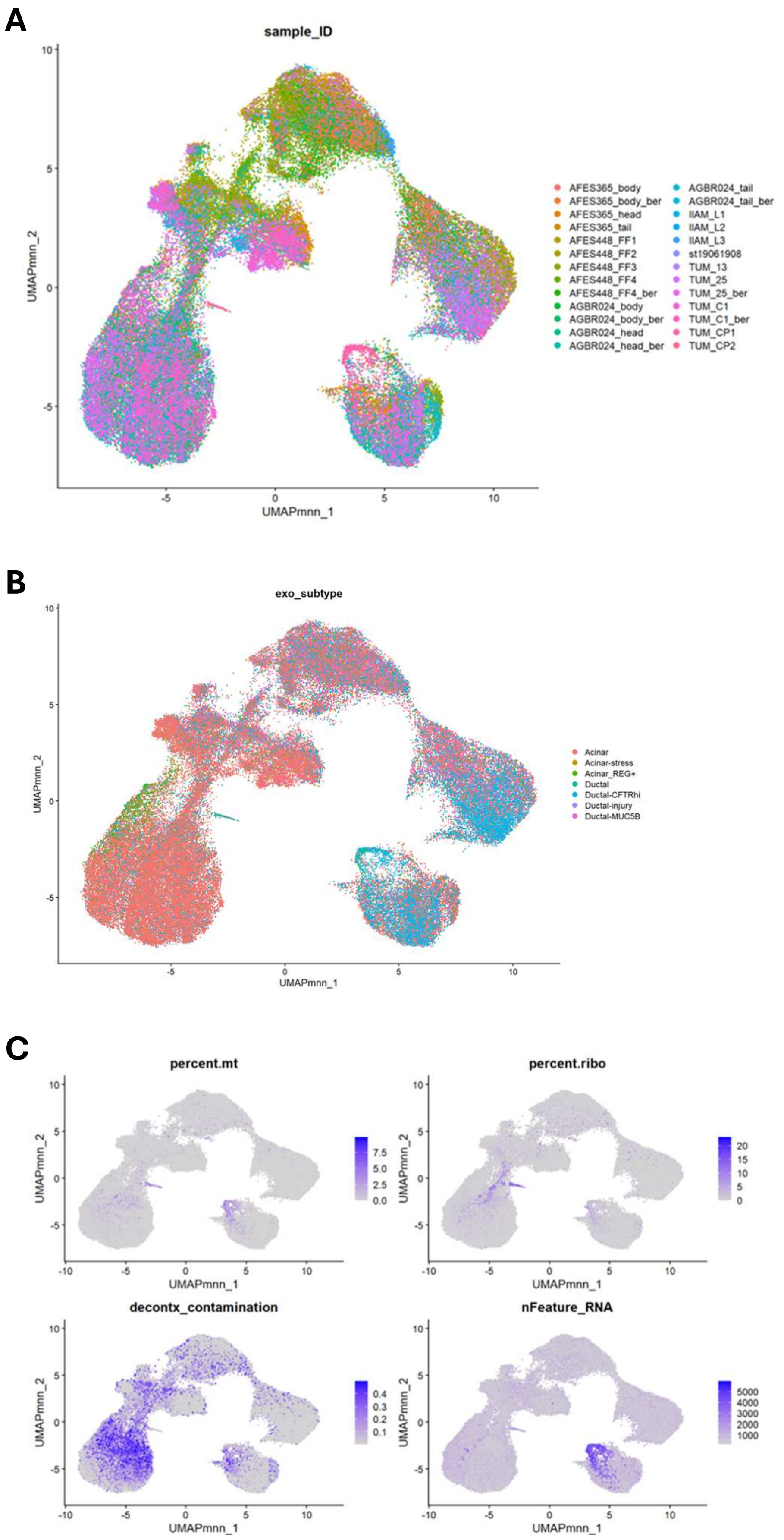


**Figure S8: FastMNN integrated exocrine cell subset of all 3 Tosti et al. (2021) datasets.** UMAP plots of exocrine pancreatic cells from FastMNN integrated neonatal, adult, and chronic pancreatitis samples shaded by sample ID **(A)**, exocrine cell subtype **(B)**, or relevant QC metrics **(C)**.

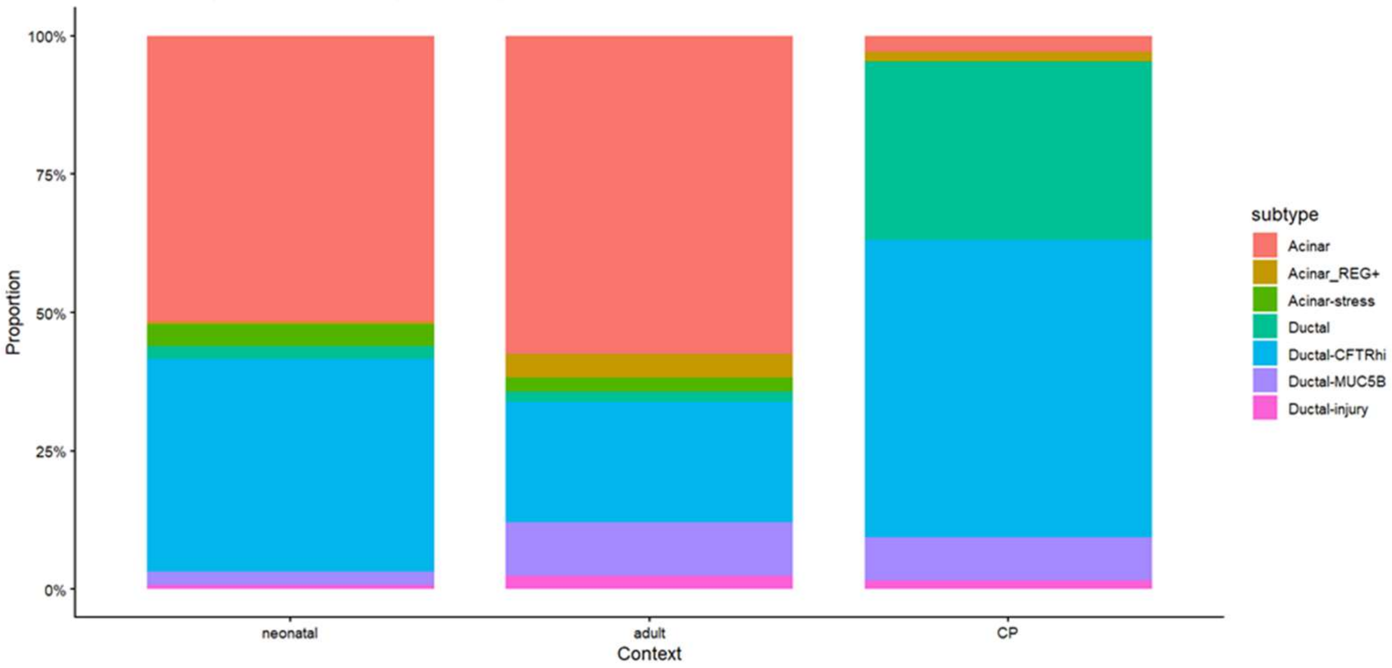


**Figure S9: Exocrine cell subtype proportions by pancreatic contexts.** Stacked bar plot indicating exocrine cell subtype proportions per pancreatic context included in the Tosti et al. (2021) snRNA-seq datasets. CP = chronic pancreatitis.

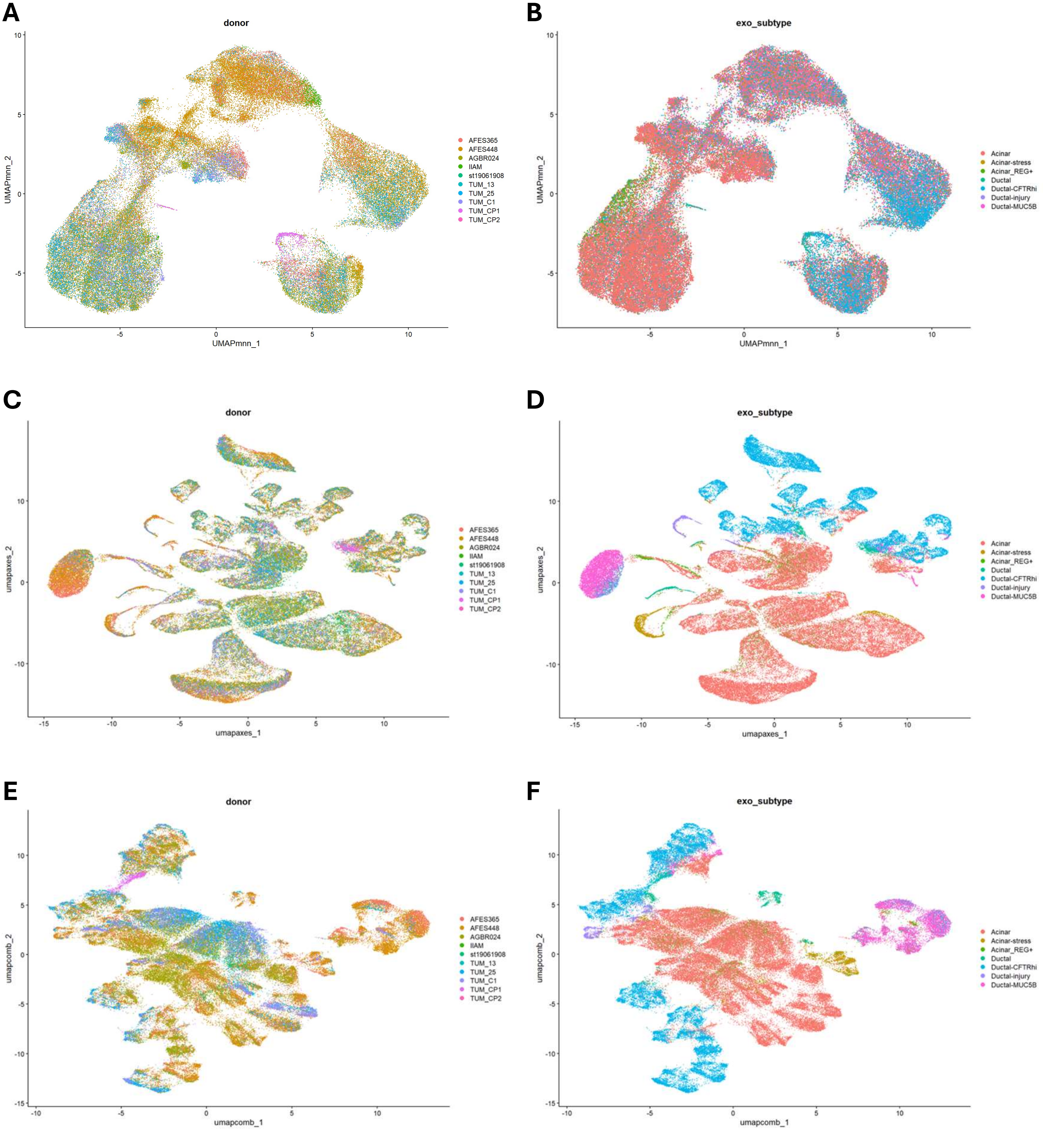


**Figure S10: MNN-based, subtype axis-based, and tuned combined embeddings.** UMAP plots of exocrine pancreatic cells with MNN-based **(A-B)**, subtype axis-based **(C-D)**, and tuned combined integration **(E-F)** of neonatal, adult, and chronic pancreatitis samples from Tosti et al. (2021) shaded by donor ID **(A, C, E)** or exocrine cell subtype **(B, D, F)**.

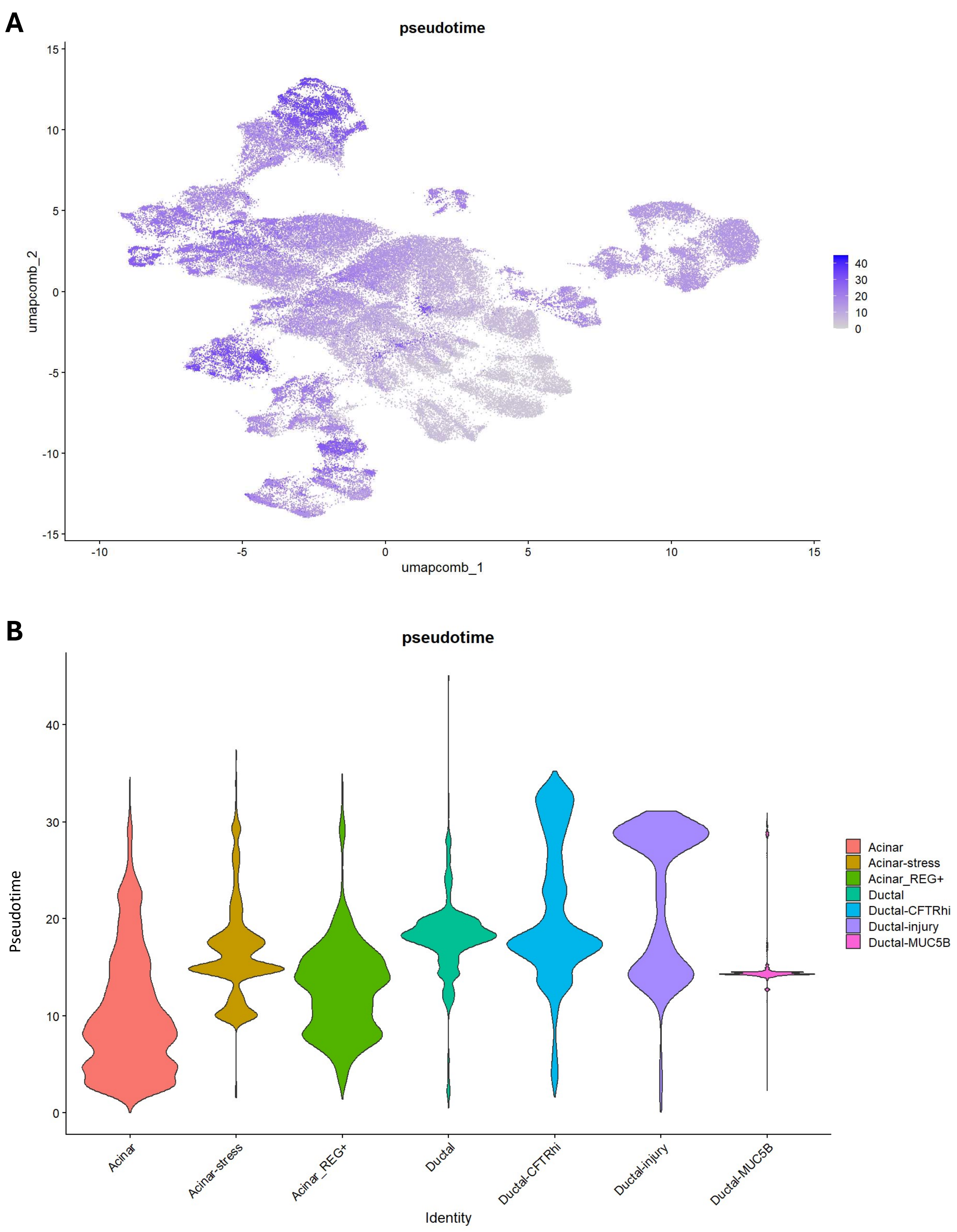


**Figure S11: Inferred pseudotime distributions across UMAP and by exocrine subtype.** **(A)** UMAP plot of exocrine pancreatic cells with tuned combined embedding shaded by Slinshot-inferred pseudotime with acinar cell subtype set as the root. **(B)** Violin plot of pseudotime distributions stratified by exocrine cell subtype.

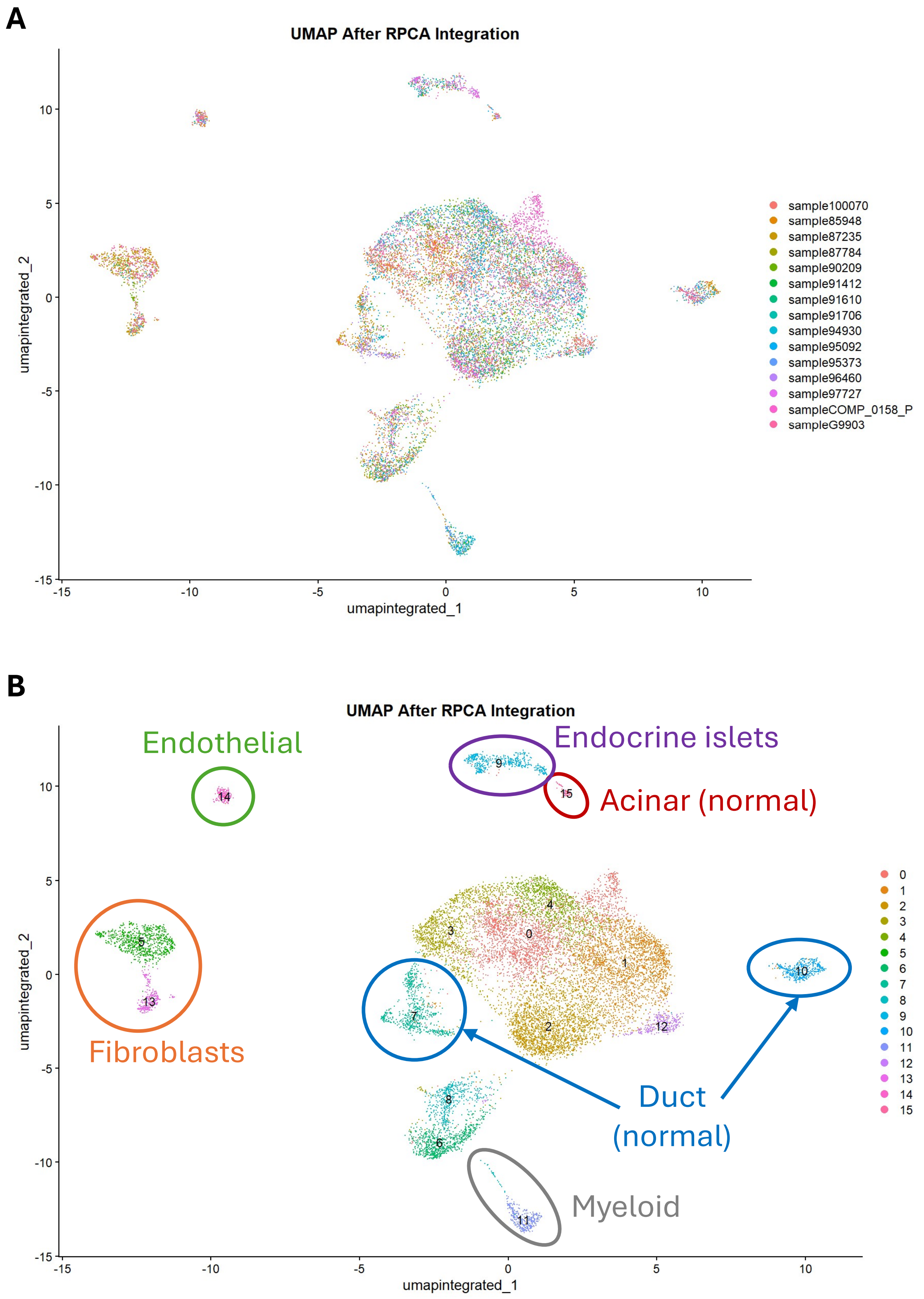


**Figure S12: RPCA integration of tumor samples and cell-type annotations.** UMAP plots of pancreatic tumor cells after RPCA integration of the Chan-Seng-Yue et al. (2020) pancreatic tumor samples shaded by sample ID **(A)** and inferred cluster **(B)**. Broad cell type annotations of non-tumor cells based on marker gene expression are indicated for inferred clusters **(B)**.
